# How threat perception of tick-borne diseases shapes preventive and control measures among dog owners in Iquitos, Peru

**DOI:** 10.64898/2026.04.28.26351956

**Authors:** Cusi Ferradas, Oliver A. Bocanegra, Winnie M. Contreras, Andrés G. Lescano, Janet Foley, Maureen Laroche

**Author notes:** **Corresponding author**: Dr. Maureen Laroche, MSc PhD FRES, Address for correspondence: The University of Texas Medical Branch, 301 University Blvd, TG Blocker MRB 4.142, Galveston, TX 77555. E.

## Abstract

Knowledge, attitudes, and practices (KAP) studies help identify priority groups for control interventions. They include multiple factors, such as threat perception—encompassing perceived susceptibility and severity of a health risk—and sociodemographic data, which have been shown to be associated with individual prevention and control measures for mosquito-borne diseases. However, their application to tick-borne diseases (TBDs) remains largely unstudied in limited-resource settings. *Ehrlichia canis*, transmitted by *Rhipicephalus sanguineus s.l.* ticks, is highly prevalent in Iquitos, Peru, a city in the Peruvian Amazon. We analyzed data from a questionnaire completed by 285 dog owners in Iquitos to assess whether the perceived threat from ticks and TBDs is linked to preventive measures for homes and dogs. Secondary aims included examining the associations between TBD awareness and preventive/control measures, as well as identifying sociodemographic factors linked to these measures and threat perception. We used simple and multiple regression models to evaluate the associations of interest. We found that 31.6% of participants (n = 90) were unaware that TBDs could occur in humans. This group tended to be older, with lower education levels and lower income. Only a small portion of the included participants reported using preventive/control measures. After adjusting for sociodemographic factors, TBD awareness was associated with a higher likelihood of home spraying. Furthermore, those who perceived TBDs as moderate diseases were marginally more likely to apply spray acaricide to their dogs, while those who perceived these diseases as severe to very severe were more likely to administer oral acaricides to their dogs. These findings highlight variability in the link between threat perception and preventive measures. Results suggest TBD awareness campaigns may benefit from focusing on older individuals with lower education and income, while educational efforts promoting effective prevention measures like acaricidal pills should target all dog owners.

**Author summary:** In Iquitos, Peru, the high prevalence of *Rhipicephalus sanguineus* s.l. infestation in dogs and homes and the high prevalence of *Ehrlichia canis* infection in dogs, highlight the need for urgent tick prevention and control. Understanding the influence of awareness, attitudes (including perceived threat) and sociodemographic factors on practices is key to designing effective interventions and identifying target groups. We analyzed data from 285 dog owners to assess if perceived threat (susceptibility and severity) from ticks and tick-borne diseases (TBDs) was linked to self-reported preventive measures for homes and dogs. We also examined TBD awareness and sociodemographic factors potentially associated with preventive practices. Among participants, 31.6% were unaware that TBDs could affect humans, with this group tending to be older and have lower education and income levels. Perceived susceptibility to tick bites and TBDs was not linked to preventive measures, while perceiving TBDs as moderate to severe increased the likelihood of using spray and oral acaricides on dogs. However, usage remained low even among those with high perceived severity. These findings reveal varied associations between threat perception and preventive practices, as they may be influenced by factors such as perceived benefits, barriers, and self-efficacy. Educational efforts should target older adults, and those with lower education and income, while campaigns promoting effective measures like acaricidal pills should address all dog owners.

## 1. Introduction

A <u>Knowledge, Attitudes, and Practices</u> (KAP) study is “*a representative study of a specific population to collect information on what is known, believed, and done in relation to a particular topic*” (World Health Organization, 2008). These surveys are valuable tools for gauging baseline knowledge levels and evaluating the impact of health education programs. They also assess attitudes, such as threat perception regarding specific health outcomes, and identify risky behaviors and potentially ineffective preventive practices. Beyond providing a descriptive overview of knowledge, attitudes, and practices, KAP surveys provide valuable data for analyzing the interplay among these factors. These studies stem from the Health Belief Model, which seeks to explain the cognitive drivers of health behavior (Champion & Sugg, 2008). This model posits that knowledge and perceived threat, which can be defined by the combination of perceived susceptibility and perceived severity of the disease, shape individual behaviors. However, it also recognizes that other factors, such as perceived barriers and whether the perceived benefits of taking preventive action outweigh the costs, influence behavior.

In the last few years, KAP studies have been increasingly used in the context of vector-borne diseases (VBDs). While most of these studies are currently focused on mosquito-borne diseases, such as malaria and dengue, KAP studies are now being extended to other arthropod vectors (De la Fuente et al., 2023; Duval et al., 2023). Ticks are the second most important arthropod vector after mosquitoes, as they are involved in the transmission of various agents, including an array of viral, bacterial, and protozoan pathogens of both veterinary and public health significance (De la Fuente et al., 2017). As with many other VBDs, vaccines against tick-borne diseases (TBDs) remain scarce. Additionally, TBDs are commonly associated with an undifferentiated febrile clinical presentation, leading to diagnostic challenges and poor patient outcomes. Therefore, it is essential for individuals to engage in preventive behaviors, such as regularly checking for ticks on themselves, their animals, and their environment, as well as using repellents, acaricides, and pesticides. KAP studies help uncover the key factors that influence the adoption of these preventive measures, offering valuable insights for improving VBD control strategies.

Most KAP studies on TBDs have been conducted in Global North* countries (De la Fuente et al., 2023), especially on Lyme disease. In the Upper Midwest of the United States, where Lyme disease is endemic, those who feel at risk of contracting the disease, those who reported themselves as very likely to get ticks, and those who considered Lyme a severe disease, were more likely to use preventive measures against ticks, indicating that perceived threat is associated with preventive practices in endemic areas (Beck et al., 2022; Niesobecki et al., 2019). In a study conducted in Ottawa, Ontario, Canada, where Lyme disease is emerging, those who perceived themselves as high risk had higher odds of having a high protective practices score (compared to those who perceived themselves as medium or low risk). However, individuals classified as having a high exposure risk (based on reported activity in woodlands during spring to fall) index, although they knew more about Lyme disease, had lower odds of having a high Lyme disease protective practices score (Logan et al., 2024). This last finding may suggest that individuals frequenting wooded areas are unlikely to encounter ticks in areas where Lyme is emerging, which may contribute to the lower use of preventive behaviors. These findings suggest that the association between threat perception and practices for TBDs may be influenced by disease endemicity. Individual factors, such as sex, ethnicity, sociodemographic status, and societal factors such as cultural norms, influence beliefs (Champion & Sugg, 2008). Therefore, results from studies conducted in Global North countries may not be generalizable to resource-limited settings, which highlights the need for local studies.

In the Peruvian Amazon basin, the high incidence of mosquito-borne diseases has made them a top priority for governmental institutions, which have focused on educational initiatives to raise awareness and promote appropriate health practices. In contrast, ectoparasite-borne diseases (*Rickettsia* spp., *Bartonella* spp., etc.), which are present at lower prevalence among humans in these areas, are yet to be the target of government prevention campaigns (Kocher et al., 2016, 2017). In urban areas like Iquitos, a landlocked city in the Peruvian Amazon, dog owners may face the risk of pathogens transmitted by *Rhipicephalus sanguineus* s.l. These ticks mainly parasitize dogs but occasionally feed on humans. Such pathogens include *Ehrlichia canis*, a vector-borne bacterium prevalent among dogs in Iquitos (Villaverde, 2017; Ferradas, 2025), which has been sporadically detected in humans in Venezuela and Italy (Sgroi et al., 2024). While *E. canis* has never been molecularly detected in humans living in Iquitos, they might be exposed to other zoonotic pathogens transmitted by *R. sanguineus* s.l. ticks, which are frequently reported in other parts of the Americas. Most notably, *Rickettsia rickettsii*, one of the most pathogenic rickettsial species, has been associated with high case-fatality rates in certain areas of Mexico and other countries in the Americas (Álvarez-Hernández et al., 2024).

The main aim of this study was to assess whether dog-owner perception of the threat posed by tick bites and TBDs to humans in Iquitos, Peru is linked to their self-reported use of preventive/control measures in both their homes and their dogs. The secondary objectives were to 1) evaluate if awareness of TBDs in humans was associated with the use of preventive/control measures for ticks, and 2) identify sociodemographic factors associated with threat perception and the use of preventive/control measures. Evaluating how threat perception drives actions against tick infestations can reveal whether raising awareness about TBDs in humans could lead to increased and improved use of preventive measures. Additionally, exploring the relationship between individual factors, attitudes, and practices can help prioritize groups for targeted educational interventions.

## 2. Methods

### 2.1. Ethical statement

This study is a secondary data analysis of a study entitled “A multidimensional analysis of the risk of infection with *Ehrlichia canis* among urban dogs in Iquitos, Peru”, nested in the umbrella study “Eco-epidemiological study of ectoparasite-borne diseases in Peru”, which was approved by the Universidad Peruana Cayetano Heredia (UPCH) IRB (N 201148) on March 13, 2020, and renewed as many times as needed until completion of the study. IACUC approval was also obtained from UPCH on April 14, 2020, and research authorizations were secured from the Gerencia Regional de Salud of Loreto (N 075-2023-GRL-GRSL/30.90) on March 28, 2023. Only dog-owners aged 18 or older who agreed to participate and signed an informed consent were enrolled in the study.

### 2.2. Study site

Iquitos is a city located in the Peruvian Amazon, in the northeast of Peru, and belongs to the department of Loreto. Its access is restricted, as it does not have roads connecting it with other cities in Peru. Transportation to the city is carried out only by air or river. Iquitos has a rainy tropical climate, with average daily temperatures ranging from a minimum of approximately 21°C to a maximum of 31°C, and an average total rainfall of 248.3 mm (World Meteorological Organization, 2024). According to data from the latest national census, the metropolitan area of Iquitos, comprising the districts of Iquitos, Punchana, Belen, and San Juan, has a total population of 411,556 inhabitants, distributed across 87,583 households. The poverty rate in these districts varies between 8.6% and 26.5%, while the illiteracy rate ranges from 2.7% to 5.6% (INEI, 2018; MIDIS, 2024).

To date, there is no official data on the canine population in the city. However, during the development of this study, it was observed that most dogs have an indoor/outdoor lifestyle and tick infestations are highly prevalent. Additionally, at least 20% of surveyed participants reported having found ticks on their bodies at some point in their lives (Ferradas, 2025).

### 2.3. Study design, home selection, and participant enrollment

This study is a secondary analysis of data collected during a primary study conducted from April 1 to April 25, 2023, therefore, our current sample size aligns with initial calculations from that study. The primary study aimed to estimate the prevalence and evaluate factors associated with *Ehrlichia canis* infection among dogs. For these aims, a minimum sample size of 286 dogs, equivalent to 286 houses (considering a conservative scenario of one dog per house), was calculated to evaluate factors potentially associated with *E. canis* infection. We chose 80% power and 0.95 confidence level and assumed a 0.2 intraclass correlation coefficient and based our calculations on a previous study conducted in Panama (Pérez-Macchi et al., 2019). Once in the field, the research team visited each randomly selected building and, if the building was not an inhabited home, if nobody was present in the home, or if no dog lived in that home, the team walked to the right until a home meeting the inclusion criteria was found.

### 2.4. Data collection

Our questionnaire was designed using previous scientific publications that thoroughly assessed knowledge, attitudes, and practices regarding TBDs in Global North countries. Questions were adjusted according to the guidance of three scientists with experience conducting human subject research and animal research in the Peruvian Amazon basin. We implemented branching logic to skip questions when the previous response rendered the next one inapplicable. We then conducted a face validation with 10 dog owners living in Iquitos to assess the clarity of questions.

The final version of the questionnaire (Sup. Material 1) covered questions on participants’ sociodemographic information, such as age, gender, education level, monthly income, and occupation, as well as their knowledge, experiences, attitudes, and practices regarding ticks and TBDs. The “<u>Knowledge</u>” section assessed participants’ understanding of tick feeding sources; awareness that dogs can contract TBD (including questions about clinical signs and vectors of *E. canis*); and awareness that humans can contract TBD (including knowledge about which ticks can transmit pathogens). “<u>Attitudes</u>” measured participants’ perception of <u>susceptibility</u> to ticks and TBDs through the questions: 1) how worried are you about the possibility of being bitten by a tick?, 2) do you think you are prone to get a TBD if you do not take preventive measures?, 3) how likely are you or someone from your family to get a TBD during next year?, 4) how worried are you about the possibility of getting TBDs?. One question was used for <u>severity</u>: how severe are TBDs in humans? Only individuals who answered “yes” to the question, “Can ticks transmit pathogens to humans?” (awareness of TBDs in humans) were asked the four questions related to threat-perception about TBDs. Those who responded with “no” or “I do not know” were not asked these questions (Figure 1). Lastly, the “<u>Practices</u>” section gathered information on two variables related to general dog care—regular veterinary visits (once per year or more) and whether the dog was neutered/spayed— and three variables related to tick prevention and control: home spraying with a “substance against ticks” (either commercially available acaricides in spray form or other perceived tick-control substances) in the last three months, the application of acaricide sprays on dogs, and the use of oral acaricides for tick prevention in dogs.

**Figure 1.**
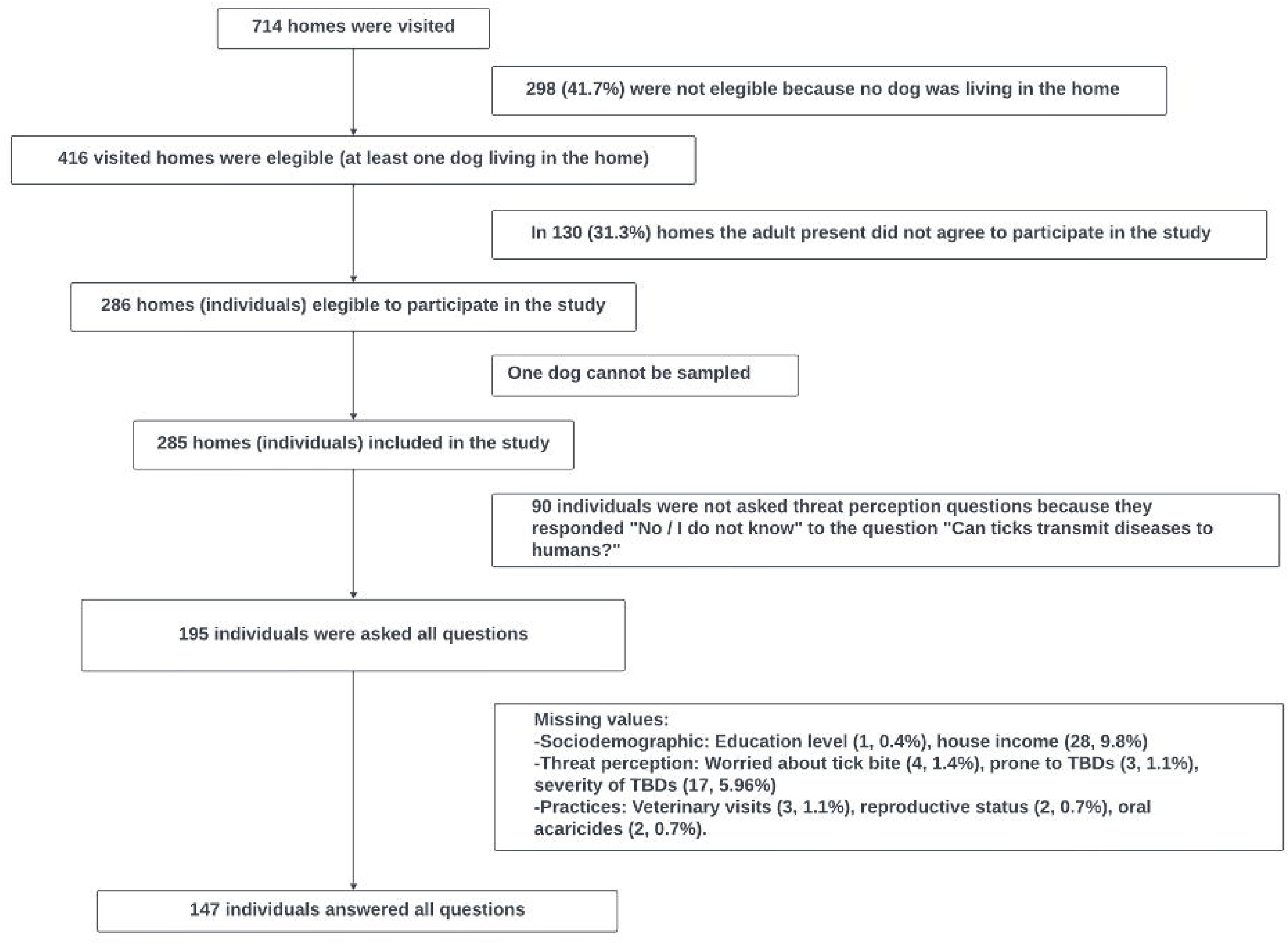
Participants selection process.

We formed two field research teams, each consisting of four members, that visited homes separately. Only one designated member from each team was responsible for administering the questionnaires during the home visits. This trained field worker verbally administered the questionnaire to all study participants, ensuring consistency and minimizing variability between interviewers. Study data were collected and managed using REDCap electronic data capture tools hosted at Universidad Peruana Cayetano Heredia. Data quality checks were performed daily by the study coordinator.

### 2.5. Variable recategorization

Sociodemographic factors included sex, age, education level, and home monthly income. Age was measured as a continuous variable but was categorized into three groups: 18–43 years (Gen Z and Millennials), 44–59 years (Generation X), and 60–80 years (Baby Boomers to Silent Generation). This categorization was selected due to its strong association with varying levels of access to information from different countries. Finally, education level was categorized as no education to attending elementary school, attending high school, attending technical education, or attending a university or graduate school. Monthly home income was recategorizes as less than S/. 1,000, between S/. 1,001 and 2,500, and more than S/. 2,500. The minimum salary in Peru is S/. 1,025. The mean wages for 2022–2023 were S/1,524.0 in Peru and S/1,581.5 in Iquitos, respectively (INEI, 2023).

All threat perception questions, except for propensity, were initially measured on a four-point Likert scale. However, they were recategorized into three categories: “worried about tick bites,” “likelihood of contracting a tick-borne disease,” and “worried about TBDs.” The two middle categories were combined, while for “severity,” the two highest categories were merged.

For the analysis of self-reported dogs’ reproductive status and administration of oral acaricides, dogs younger than 6 months and 2.5 months, respectively, were excluded, as these practices are not generally recommended before those ages.

For all self-reported tick prevention and elimination measures in dogs, we dichotomized the responses into “not used” and “used in at least one dog” to account for homes with one or two dogs. We assumed that if the owner implemented the measure for at least one dog, it indicated their awareness of its importance.

### 2.6. Statistical analysis

Analyses were conducted to separately examine four sets of relationships: (1) the association between each measure of threat perception (exposures) and each self-reported tick prevention/control measures (outcomes), (2) the association between awareness of TBDs in humans (exposure) and preventive/control measures, (3) the association between sociodemographic factors (exposures) and threat perception measures, and (4) the association between sociodemographic factors (exposures) and self-reported tick prevention/control practices (outcomes).

Chi-square or Fisher’s exact tests (two-sided) were used to compare the relative frequencies of each category of the exposure variables with the outcome variables. Then, simple generalized linear regression models with a binomial or Poisson (due to non-convergence of binomial) family, log link, and robust variance were used to assess the relationships between threat perception, awareness of TBDs in humans, and sociodemographic factors with self-reported practices. This approach calculated prevalence ratios (PR). To assess the relationship between sociodemographic factors and threat perception, multinomial logistic regression was applied to estimate odds ratios (OR).

Multiple regression analysis was conducted using a Poisson family with a log link and robust variance to evaluate the association between threat perception and awareness of TBDs in humans, adjusting for sociodemographic factors, and considering the use of preventive/control measures. Only threat perception variables that were significantly associated with at least one preventive/control measure in the simple regression analyses were analyzed using multiple regression. Additionally, we performed a third group of multiple regression models to investigate which sociodemographic variables independently affect the use of preventive/control measures without considering awareness of TBDs or threat perception of ticks and TBDs.

Missing responses for covariates (if they represented less than 2% of the observations) or outcomes of interest were excluded from regression models to ensure the completeness of data for each model. Associations with a p-value of less than 0.05 were considered statistically significant. All statistical analyses were conducted in Stata 15.0 (Stata Corp., College Station, TX)

## 3. Results

### 3.1. Sociodemographic characteristics and TBD threat perception levels of study participants

A total of 285 individuals were included in the study, comprising 196 women (68.8%). Most individuals were younger than 43, with Generation Z (born mid-1990s to early 2010s) being the largest group (54.39%). Participants had varied levels of education, ranging from no formal education to graduate school, with the largest group having completed high school as their highest level of education (35.8%). Only 89 (29.82%) lived in homes with a total monthly income exceeding S/2,500 (Table 1).

**Table 1.**
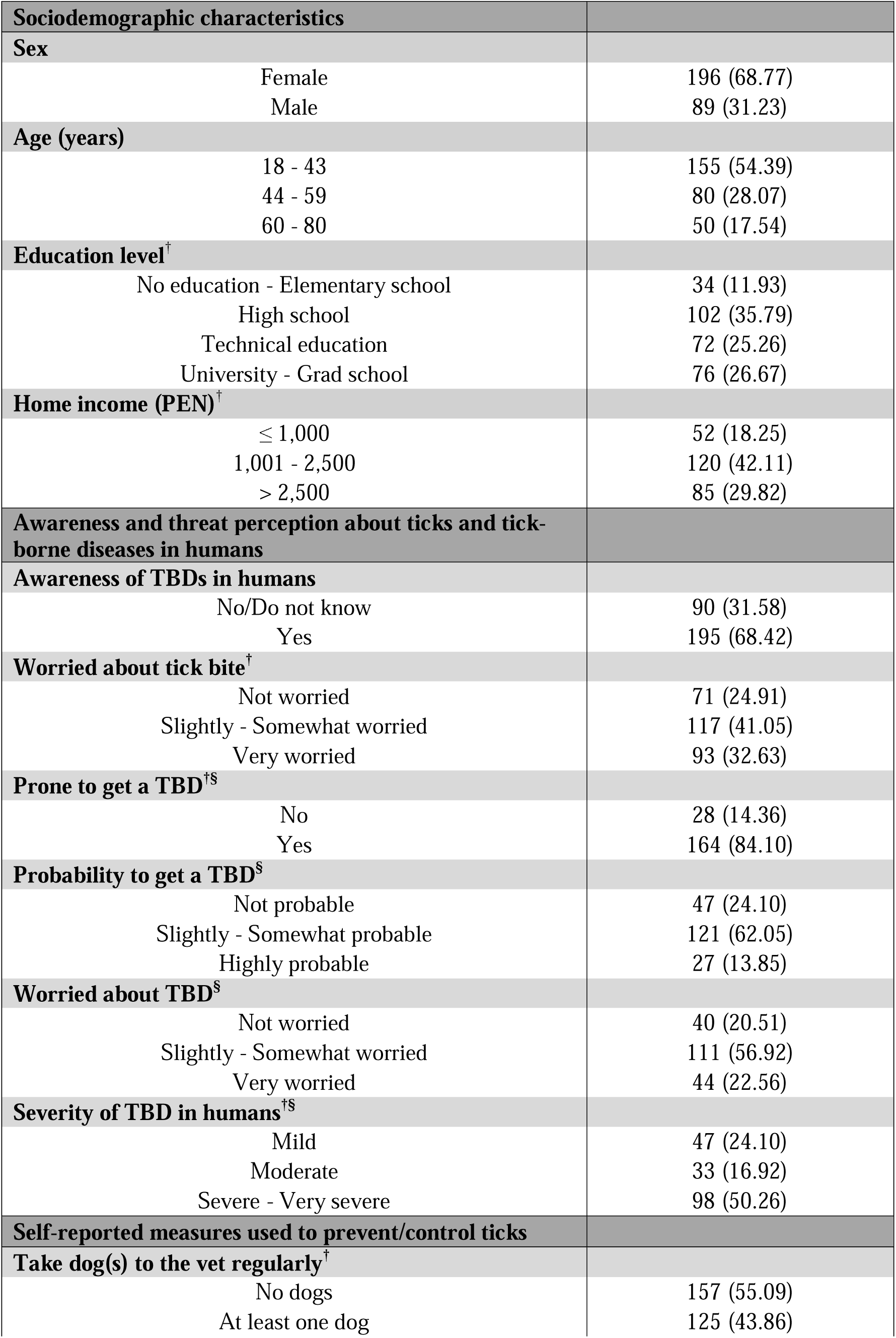
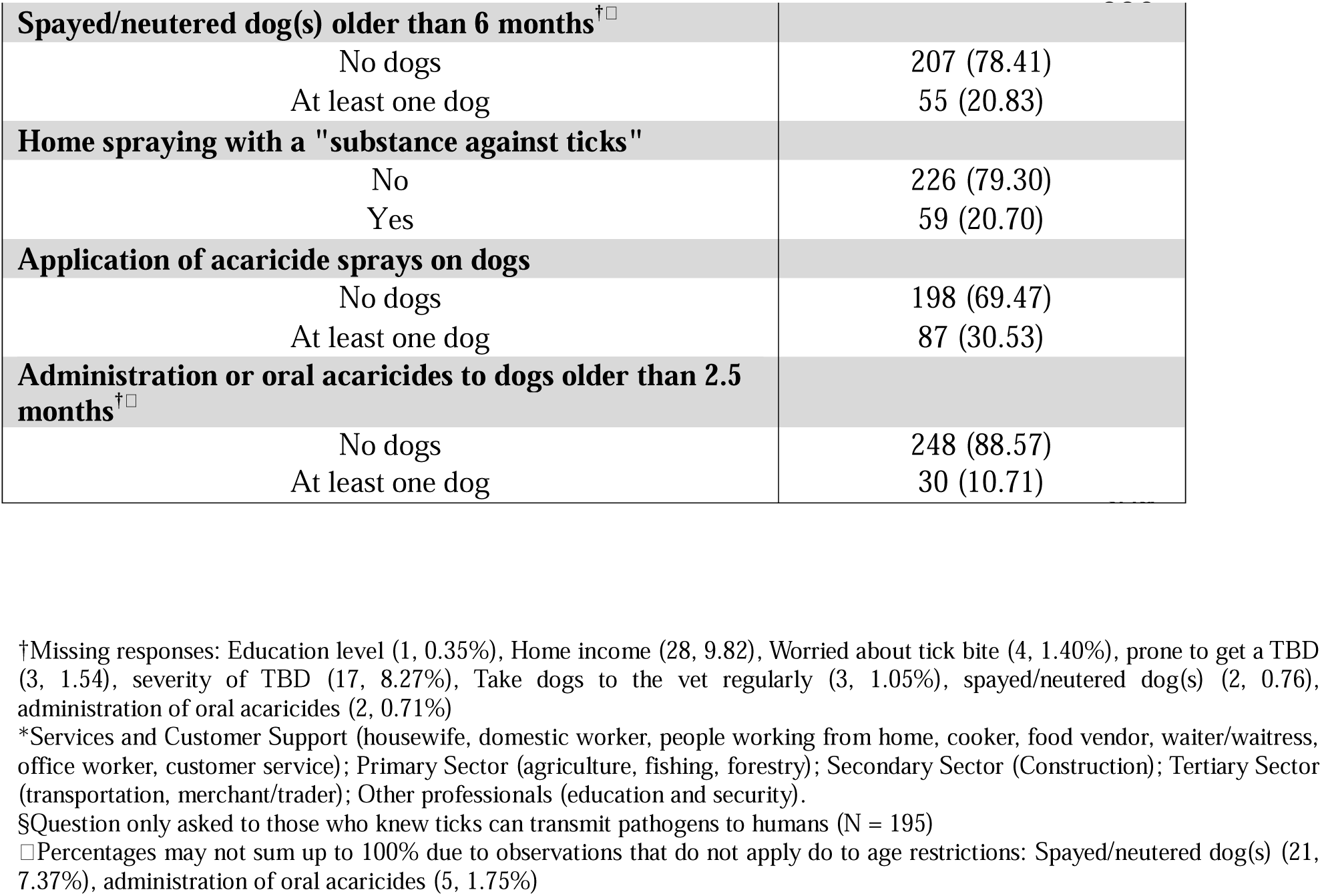
Characteristics of dog owners living in Iquitos, Peru, awareness levels of tick-borne diseases and self-reported vector control methods.

Of the 285 individuals included in the study, 93 (32.63%) indicated being “very worried” about tick bites, despite 195 (68.42%) being aware that ticks can transmit human pathogens. Among these, 164 (84.10%) considered themselves likely to contract TBDs if they did not take preventive measures, yet, only 27 (13.85%) believed it was “highly probable” that they or a family member would get a TBD in the following year. While 111 (56.92%) reported being “slightly to somewhat worried” about potentially contracting a TBD, only 44 (22.56%) were “very worried” about these diseases, despite 98 (50.26%) considering them to be “severe to very severe” among humans (Table 1).

### 3.2. Self-reported measures to prevent/control ticks

A total of 125 individuals (43.86%) reported taking their dog(s) to the veterinarian regularly, most of them (n=53; 42.40%) every 2-3 months. However, only 55 (20.83%) had neutered or spayed their dog(s) older than six months.

Acaricide use was moderately low with 59 individuals (20.70%) indicating they spray their homes with a “substance against ticks,” 87 (30.53%) apply spray acaricides to their dog(s), and 30 (10.71%) administer oral acaricides to their dog(s) older than 2.5 months (Table 1). Most participants (n=25; 42.37%) reported using commonly available substances for home spraying, such as pyrethroids and phenylpyrazoles, while others indicated using phenol-based products, including Creoline, petroleum (sometimes mixed with bleach), endectocides and ectoparasiticides like ivermectin and amitraz, or disinfectants, mainly bleach. Thirteen participants (22.03%) did not recall the names of the products used, although four of them (30.77%) noted using “an acaricide powder diluted in water” (Sup. Table 1). Most participants who used spray acaricides to treat their dogs against ticks relied on commercially available products, mainly Fipronil-based formulations, reported by 64 participants (73.53%). However, 23 (26.47%) could not recall the name. Lastly, of the 30 participants administering oral acaricides to their dogs, 23 (76.67%) identified specific products, such as Bravecto, Simparica, Nexgard, and Atrevia, while seven (23.33%) did not recall the names.

### 3.3. Assessing the relationship between threat perception and preventive/control measures

Among individuals who reported being aware of TBDs in humans, only the perceived severity of TBDs was associated with the use of some preventive or control measures in the simple regression models. Those who perceived TBDs as moderate or severe to highly severe were more likely to take their dogs to the veterinarian regularly than those who perceived TBDs as mild. Additionally, those who perceived TBDs as severe/very severe were marginally significantly more likely to administer oral acaricides to their dogs (Table 2 & Sup. Table 2).

**Table 2.**
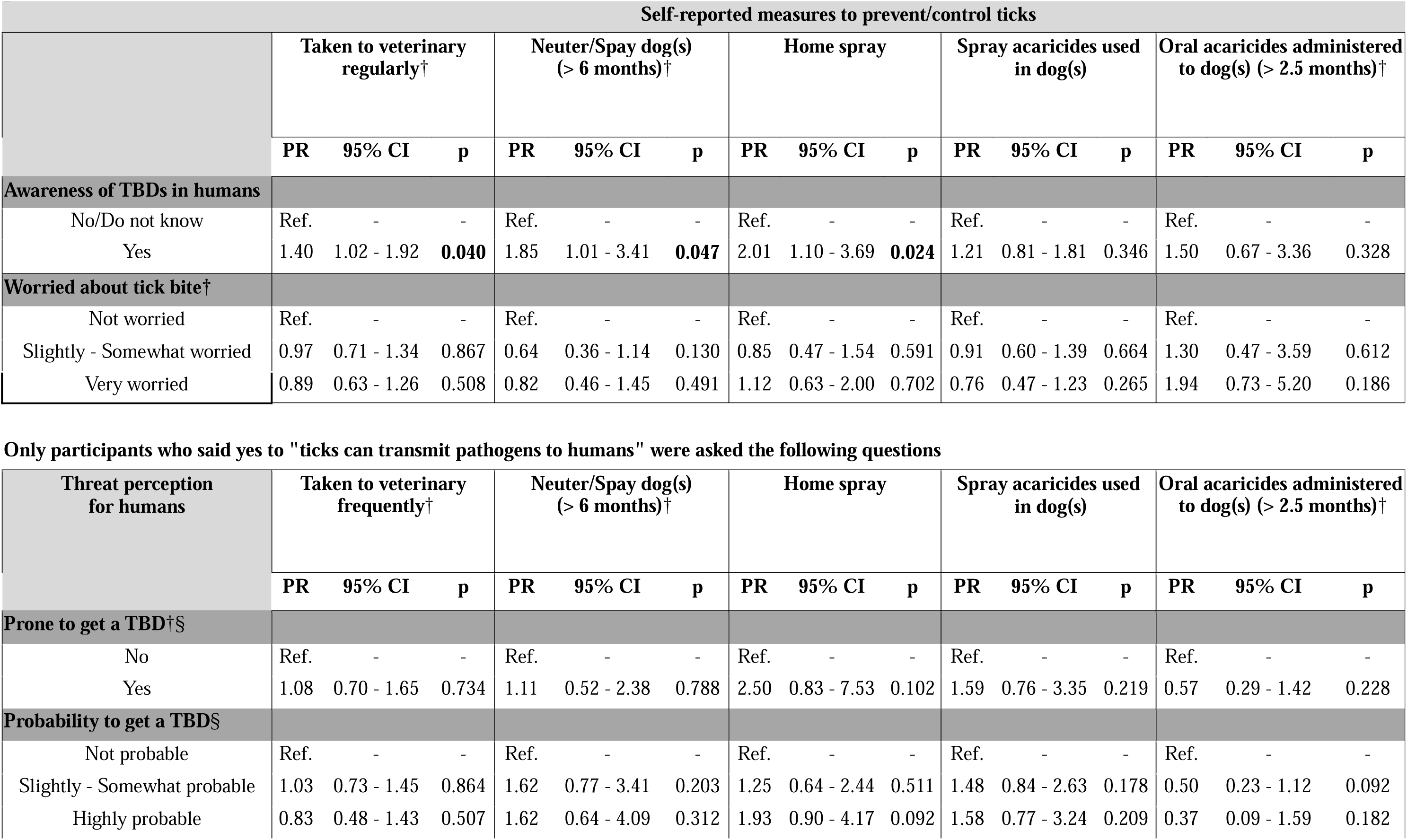
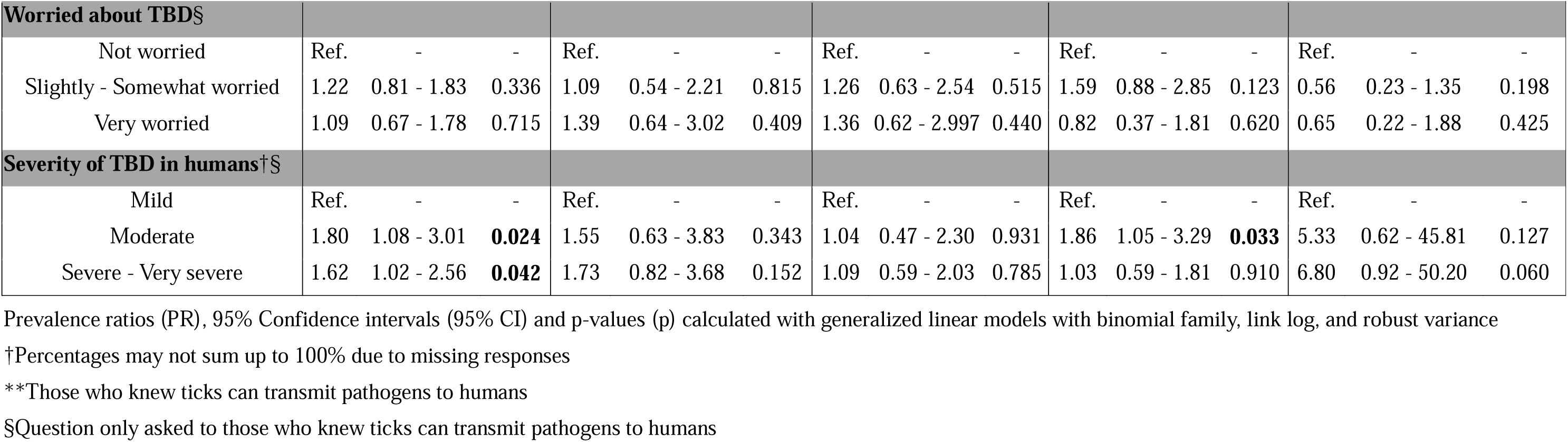
Simple regressions between awareness of TBDs and threat perception for humans with self-reported measures to prevent/control ticks.

### 3.4. Assessing the relationship between awareness of TBDs in humans and preventive/control measures

In the simple regression, individuals who reported being aware of TBDs in humans were more likely to take their dog(s) to the veterinarian regularly and to have their dog(s) spayed or neutered. They were also more likely to spray their homes with “substances against ticks” (Table 2 & Sup. Table 2). However, after adjusting for sociodemographic variables, knowing that ticks can transmit pathogens to humans was only statistically significantly associated with the use of home sprays (PR 1.87, 95% CI 1.01–3.47) (Table 3).

**Table 3.**
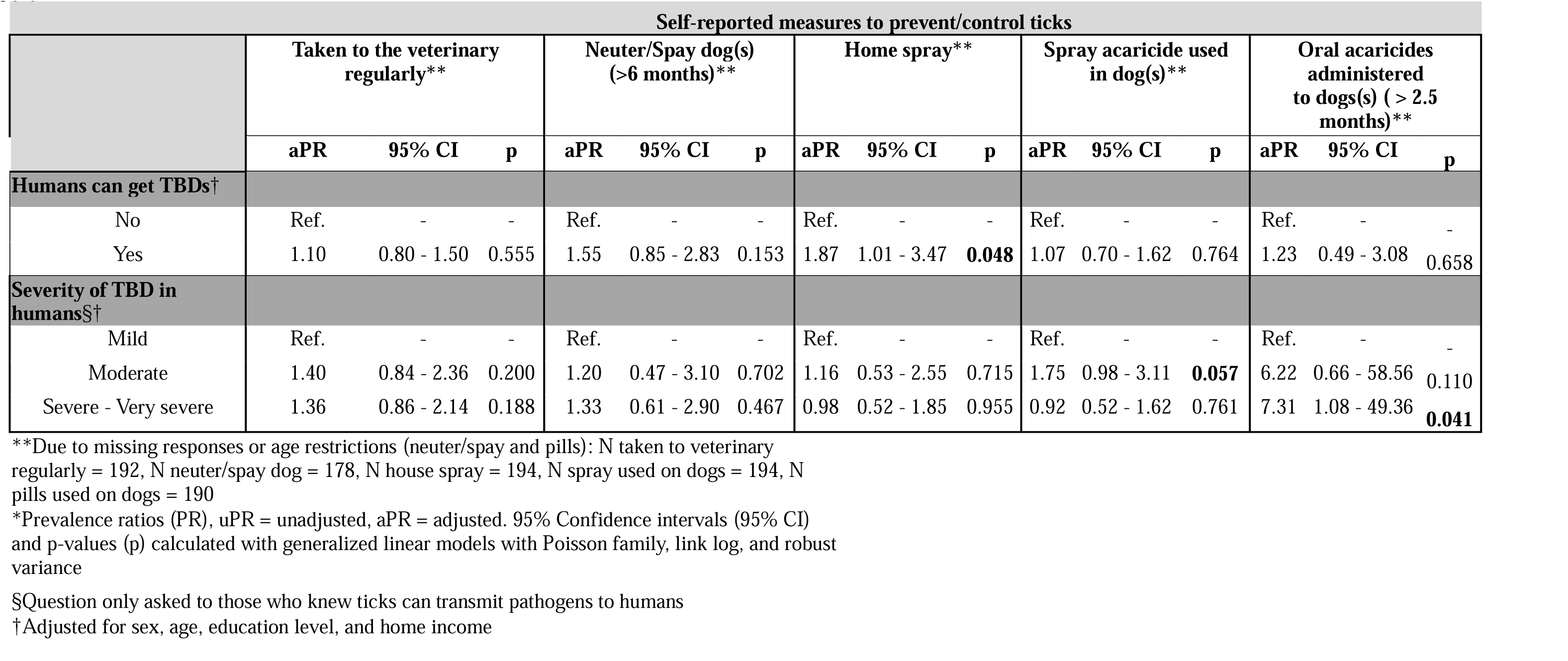
Association between awareness of TBDs or perceived severity for humans and self-reported measures to prevent/control ticks using a multiple regression analysis adjusted for socioeconomic factors.

### 3.5. Assessing the relationship between sociodemographic factors and preventive/control measures

In simple regression models, several sociodemographic factors were found to be positively or negatively correlated with veterinary care for dogs. However, after adjusting for other sociodemographic variables, we found that men were less likely to take their dogs to the veterinary frequently (PR 0.73, 95% CI 0.54–0.99), while people older than 60 (PR 1.73, 95% CI 1.21–2.49) and those with higher education (PR 3.47, 95% CI 1.78-6.76) were more likely to take their dogs to the veterinary regularly. Details are presented in Table 4.

**Table 4.**
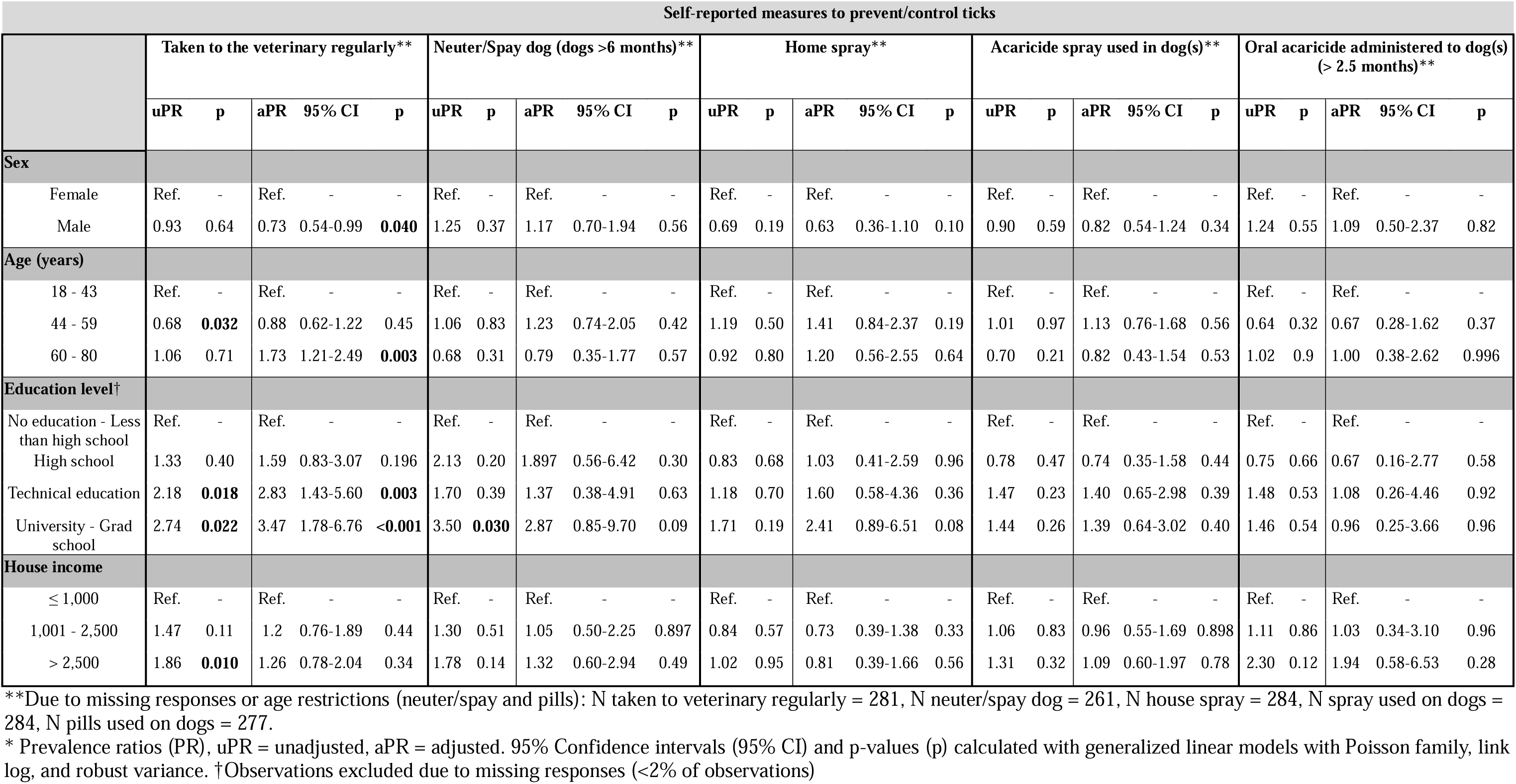
Associations between sociodemographic factors and self-reported measures to prevent/control ticks using simple and multiple regression analyses.

### 3.6. Assessing the relationship between sociodemographic factors and threat perception

Men and people over 44 were found to have lower odds of worrying about tick bites, in congruence with older groups showing lower odds of knowing that ticks can transmit pathogens to humans.

Individuals with higher levels of education had higher odds of knowing that ticks can transmit pathogens to humans and perceiving TBD as highly severe. Yet, those who stopped their education after high school had higher odds of recognizing “slightly to somewhat probable” that they or a family member would get a TBD in the next year if they did not take preventive measures.

Finally, individuals living in a home with a monthly income of more than S/. 2,500 had higher odds of knowing that ticks can transmit pathogens to humans. Additionally, among those who knew, individuals in the highest income category also had higher odds of only being “slightly to somewhat worried” about TBDs (Table 5a, Table 5b, & Sup. Table 2).

**Table 5a.**
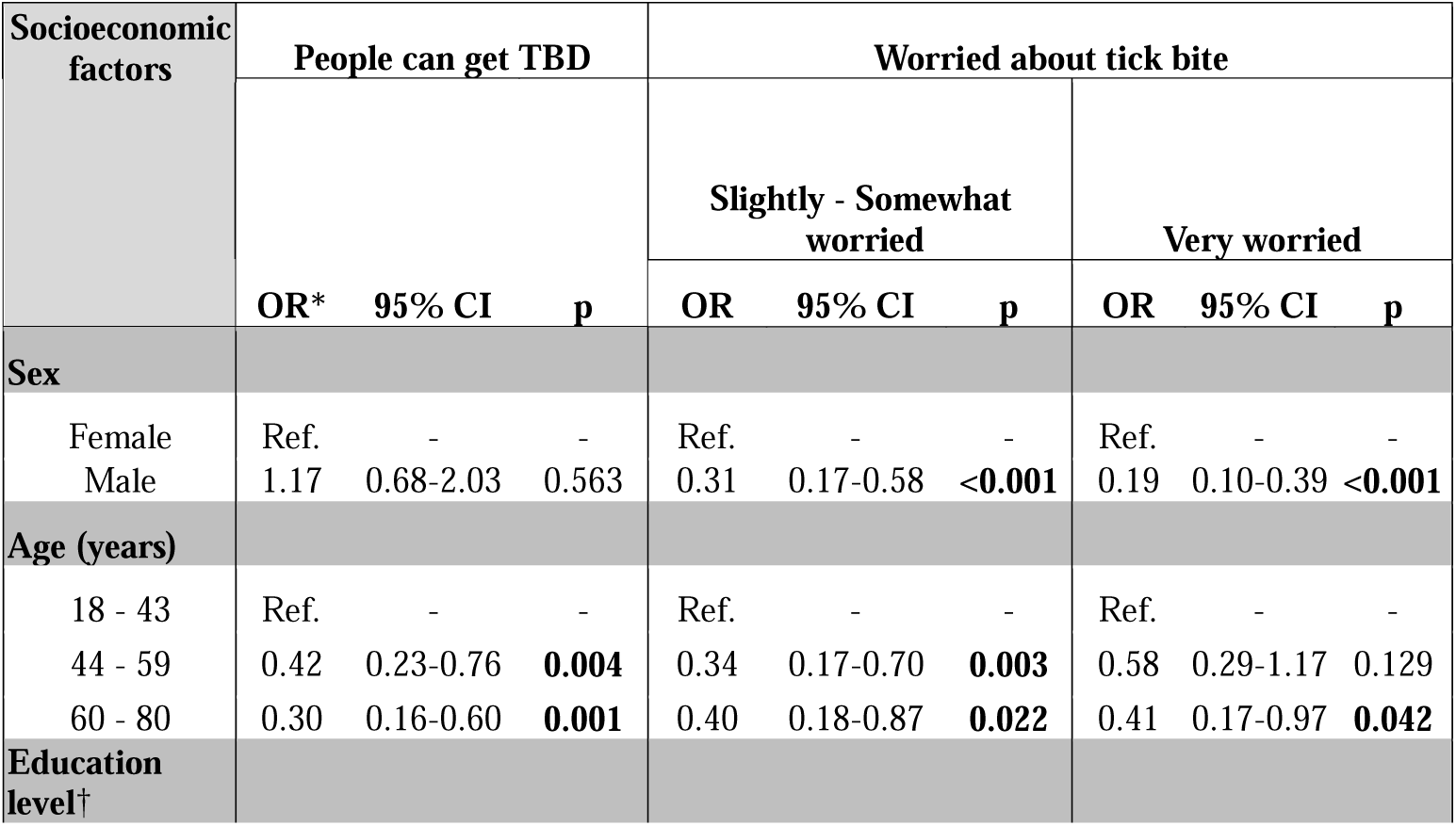
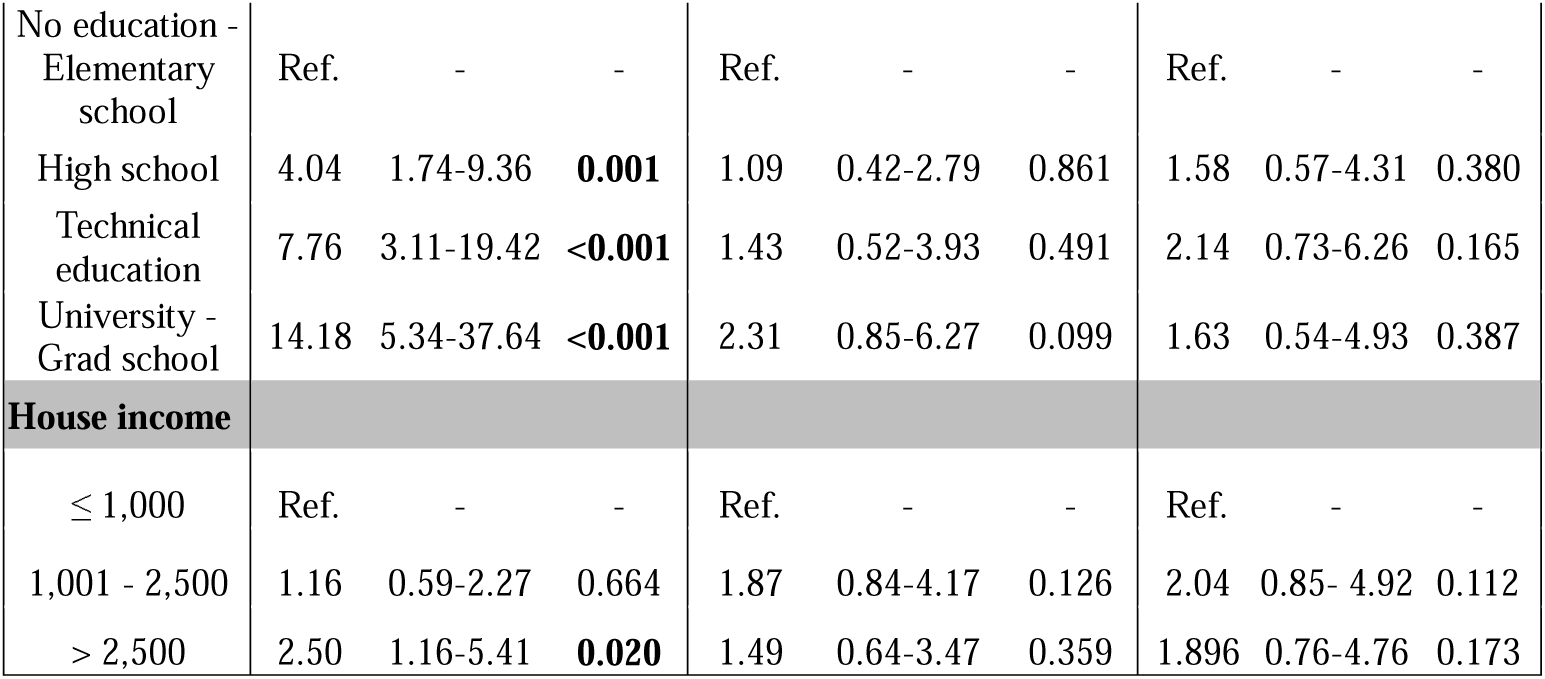
Sociodemographic information of all participants assessed for awareness of TBDs or threat perception.

**Table 5b.**
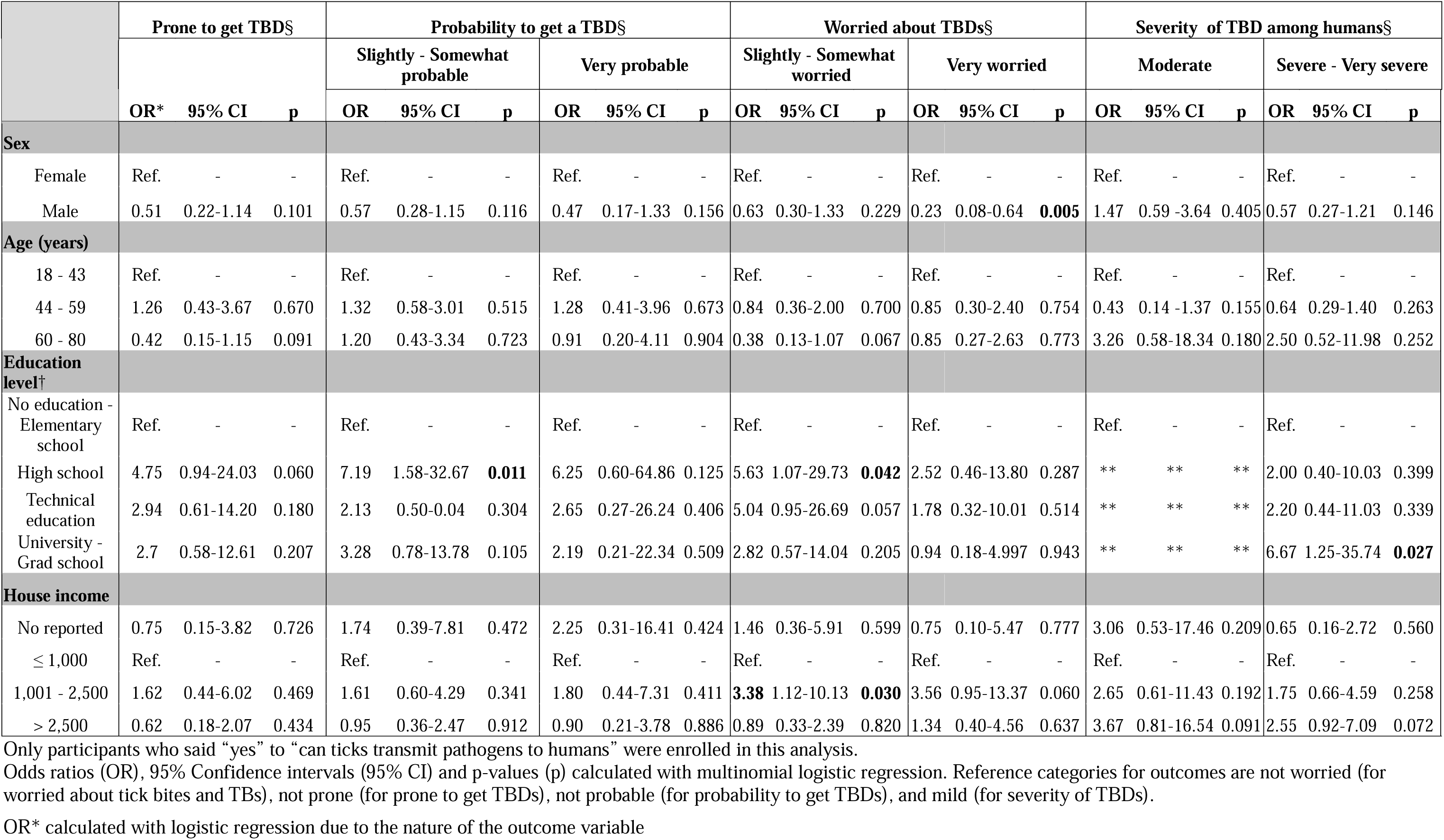
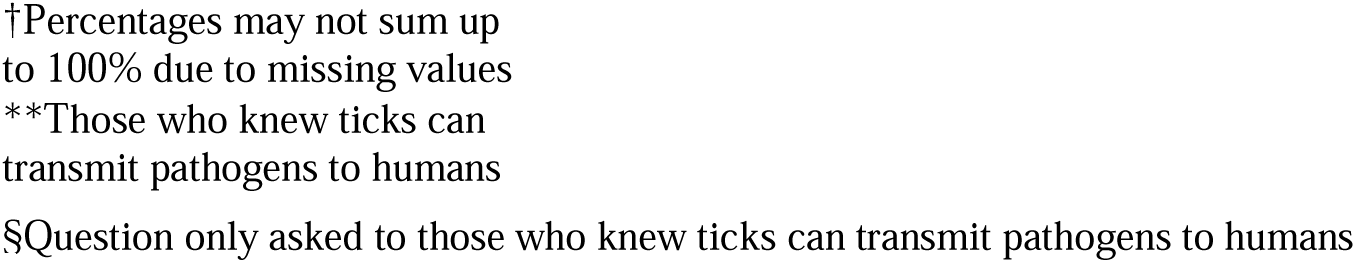
Associations between sociodemographic factors and awareness of TBDs or threat perception using simple regression analyses.

## 4. Discussion

We conducted a study with the primary aim of assessing whether dog owners’ perception of the threat posed by tick bites and TBDs to humans in Iquitos, Peru, was linked to their self-reported use of preventive and control measures in both their homes and their dogs. The threat perception of TBDs was assessed among participants who were aware that ticks could transmit pathogens to humans (68.42%). Indeed, almost 32% of participants were unaware that ticks can transmit pathogens to humans, underscoring the need to prioritize awareness campaigns about the health risks associated with ticks. In contrast, only 2% of individuals in the U.S. Upper Midwest, where Lyme disease is highly endemic, were unaware of the disease (Beck et al., 2022). Among our participants who recognized the risk of TBDs in humans, the use of specific prevention/control measures depended on the perceived severity of the diseases, but not the perceived susceptibility to them, indicating variability in how threat perception influences preventive and control behaviors for ticks among individuals in Iquitos.

Perceived severity of TBDs was linked to an increase in effective preventive measures for dogs, including sprays and oral acaricides. However, caution is warranted in interpreting the association with oral acaricides due to a wide confidence interval. Some individuals who perceive TBDs as severe may have experienced a dog with symptomatic *E. canis* infection, which could heighten their fear of TBDs and awareness of effective tick prevention measures. However, understanding the relationship between perceived threat perception and the adoption of preventive/control practices is not always straightforward, as it may be influenced by a range of individual beliefs, including confidence in preventive/control measures and perceived benefits (Champion & Sugg, 2008; Muleme et al., 2017; van Doorn et al., 2007). The lack of association between proxies of perceived susceptibility (such as concern about tick bites, probability of getting TBDs, and worry about TBDs) and the use of preventive measures was unexpected. It may relate to the belief that these preventive actions will not work, though this construct was not measured in the present study (Beck et al., 2022). Owned dogs sometimes still acquire ticks despite their owners’ attempts to use acaricides, leading to a perception of being unable to prevent vectors and VBDs. Additionally, the lack of association between perceived susceptibility and the use of preventive/control measures may stem from the fact that most of our study population fell into the “slightly to somewhat worried” and “slightly to somewhat probable” categories regarding the likelihood and concern about TBDs. Findings from previous studies suggest that moderate levels of threat perception do not have the same impact as extreme levels. At moderate levels, the perceived benefits of preventive/control measures may not outweigh the perceived costs (van Doorn et al., 2007). However, this does not completely align with our findings, as moderate severity was a significant predictor of acaricide spraying on dogs.

Patients with are rarely tested for TBS, even with compatible clinical signs and after testing negative for other pathogens (Kocher et al., 2016), which was shown to be associated with reduced awareness in the context of mosquito-borne diseases (Duval et al., 2023; Fonzo et al., 2024). However, the level of awareness of TBDs in our study population (≈ 68%) may be due to the high prevalence of tick infestations and tick-borne pathogens like *E. canis* and possibly *Anaplasma platys*, which often cause visible disease in dogs (Namgyal et al., 2021), thus potentially contributing to a heightened sense of threat in dog owners.

Importantly, the associations between awareness of TBDs in humans and preventive measures (such as regular veterinary visits and spaying/neutering) lost strength in the adjusted models, suggesting that sociodemographic variables confound these associations. Those aware of TBDs in humans tended to be younger, more educated, and had higher incomes, variables that were also associated with dogs’ general care. The association between higher socioeconomic status and awareness of TBDs has been previously reported (Reyes-Castro et al., 2021). The association between awareness of TBDs in humans and home spraying remained significant in the multiple regression models. Home spraying, in particular, may reflect the perception that seeing ticks in their home environment poses a greater risk to humans than observing them on dogs. Moreover, home spraying is less expensive than spraying acaricide on dogs. The lack of association between awareness of TBDs and spaying/neutering dogs may also be related to the fact that this practice is likely viewed strictly as a method to prevent dog pregnancies and may not be perceived as contributing to a reduction in dog roaming, and therefore, in tick infestation and tick-borne disease burden. However, we considered it an essential practice of general dog care as it may be associated with dog grouping behaviors where male *R. sanguineus* s.l. can pass from one dog to another. Additionally, populations of dogs with high percentages of reproductively intact dogs lead to the constant production of immune-naïve puppies for TBDs.

As expected, sociodemographic predictors of regular visits to a veterinary clinic included sex, age, and higher education levels. Education level is a key factor influencing the appropriate use of preventive practices to prevent mosquito-borne diseases (Arellano et al., 2015; Bohmann et al., 2022; Mouchtouri et al., 2017; Raude et al., 2012; Tuiten et al., 2009; Walker et al., 2018), including in Iquitos, where dengue prevention campaigns are constantly conducted (Paz-Soldán et al., 2015). In our study, higher education was not associated with practices to prevent/control ticks on dogs, suggesting that educational campaigns focused on these practices should include all people in Iquitos, regardless of education level.

Women were more likely than men to take their dogs to the veterinarian, possibly in part because they also expressed greater concern about tick bites and TBDs. Previous research has shown that women tend to be more concerned about health issues (Bahrami & Yousefi, 2011; Robichaud et al., 2003). Some of the concerns may arise because *Rhipicephalus sanguineus* s.l. is endophilic and women in our target population typically spend more time indoors compared to men (Dantas-Torres, 2010; Weldon et al., 2018). Individuals between 60 and 80 years old were less likely to express high levels of concern about tick bites. This may be attributed to life experiences (i.e., reaching an old age without having experienced a severe TBD) or other age-related factors, such as a reduced ability to notice tick bites due to uneven skin pigmentation from aging keratinocytes, peripheral neuropathy, and declining vision. In contrast, younger individuals aged 18 to 43 may have greater access to information (e.g., at school, on social media) and fewer age-related issues that impact their ability to notice ticks. Although we did not collect this specific information, one participant shared that she learned about tick-borne diseases from seeing on TikTok that “the singer Thalía shared her Lyme disease diagnosis.”

Our study has several limitations. First, as a cross-sectional study, we could only identify associations without establishing causality. We recognize that threat perception and practices are interconnected and impact one another. Indeed, while threat perception impacts the use of preventive/control measures, these measures likely impact threat perception levels in turn. Additionally, we did not ascertain whether the interviewed individual was primarily responsible for the dog’s care, which could have influenced the associations assessed. However, it is reasonable that the characteristics and threat perceptions of all home members collectively influence the dog’s care, thereby supporting the validity of our findings. Furthermore, we relied on self-reported practices. To mitigate response bias, we asked for the names of the products used, their frequency of use, and methods of application (Zarei et al., 2024). Although these responses can sometimes be validated using veterinary records in other contexts, access to such records is frequently restricted in low-income settings, where digital systems are unavailable and veterinary records are inconsistently kept by owners. Ultimately, due to the observed variability in associations between threat perception and proxies for preventive and control measures, we opted not to create composite scores for these constructs. Consequently, we could not determine whether higher perceived threat corresponds with greater use of preventive/control measures, nor assess whether this relationship might be nonlinear, as reported for KAP components in prior studies (Muleme et al., 2017; van Doorn et al., 2007).

Future studies should use methods such as focus groups to gain a deeper understanding of threat perception, self-efficacy, perceived benefits, and barriers to preventing and controlling tick infestations in dogs and homes. Comparing the perceived susceptibility and severity of TBDs to highly prevalent VBDs, such as dengue and malaria, could provide valuable insights, as has been done in previous research (Weldon et al., 2018). Additionally, it is essential to explore the willingness to use acaricides in both dogs and home environments, along with the perceived risks associated with these products. Researchers should also investigate preferred informational resources such as lectures, printable materials, social media, or others. Furthermore, additional studies should assess KAP methods within a Global South context, where confirming self-reported use of preventive measures and seeking medical care is particularly challenging. Additionally, while willingness to pay has been identified as a strong predictor of willingness to adopt tick prevention measures in the Global North, its applicability in the Global South is more complex. This is because individuals who express a willingness to pay may lack the financial means to follow through (Enriquez et al., 2024).

While this study was a secondary analysis of data collected for a primary study focused on *E. canis*, it provides valuable insights on threat perception in the context of tick-borne diseases in general, especially diseases associated with the cosmopolitan *Rhipicephalus sanguineus* ticks. In resource-limited settings, the success of public health interventions relies heavily on the understanding of socioeconomic factors, as well as the capacity of individuals and communities to recognize disease-related risks and implement preventive measures (Muleme et al., 2017). KAP surveys, therefore, provide critical insights to tailor interventions to the specific needs, perceptions, and barriers within a population, ultimately enhancing the effectiveness and sustainability of health initiatives. Our findings indicate that educational strategies should take into account the diverse composition of the population (Reyes-Castro et al., 2021). They may benefit from prioritizing older individuals with lower education and lower home income in campaigns to raise awareness about TBDs in humans and communicating effective measures for managing ticks, both in the environment and on dogs, targeting all education levels.

## Data Availability

All data produced in the present work are contained in the manuscript and its supplementary files.

## 5. Acknowledgements

Authors would like to thank our participants, as well as the field team (Brandon Moreno, Magno Zambrano, Carolina Ojeda, Ana del Aguila, Estefany López, Sandra Chávez, and Arturo Chávez) for their dedicated work. Additionally, we would like to thank Dr. Amy Morrison for providing the Iquitos basemap and Román Llontop for his assistance in translating the study questionnaire into English for publication.

## 8. Definitions

In this manuscript, inequalities in research are reflected in the use of Global South/Global North countries. Although these terms are imperfect and erase individual national strategies, they reflect, with no negative connotation, disparities in access to healthcare, resources, and more. In the context of academia, these differences encompass access to research grants, international conferences, internships and collaborations, research facilities, high-impact journals, and other opportunities.

Additionally, “S/.” denotes *soles*, the official currency of Peru.

## 10. Supporting information caption

**Sup. Table 1.** Type of products used by the participants to treat their house against arthropods.

**Sup. Table 2.** Associations between awareness of TBDs or threat perception for humans and self-reported measures used to prevent/control ticks using a bivariate analysis.

**Sup. Table 3.** Associations between sociodemographic factors and self-reported measures to prevent/control ticks using bivariate analyses.

**Sup. Table 4a.** Associations between sociodemographic factors and threat perception for humans using bivariate analyses.

**Sup. Table 4b.** Associations between sociodemographic factors and threat perception for humans using bivariate analyses in participants who said yes to “ticks can transmit pathogens to humans”.

**Sup. Table 5.** Characteristics of individuals excluded from final analyses due to missing values for home income and/or perceived TBD severity.

