## Supplementary material for "How threat perception of tick-borne diseases shapes preventive and control measures among dog owners in Iquitos, Peru": Table 1

|  | **N (%)** |
| --- | --- |
| **Sociodemographic characteristics** |  |
| **Sex** |  |
| Female | 196 (68.77) |
| Male | 89 (31.23) |
| **Age (years)** |  |
| 18 - 43 | 155 (54.39) |
| 44 - 59 | 80 (28.07) |
| 60 - 80 | 50 (17.54) |
| **Education level**^†^ |  |
| No education - Elementary school | 34 (11.93) |
| High school | 102 (35.79) |
| Technical education | 72 (25.26) |
| University - Grad school | 76 (26.67) |
| **Home income (PEN)**^†^ |  |
| ≤ 1,000 | 52 (18.25) |
| 1,001 - 2,500 | 120 (42.11) |
| > 2,500 | 85 (29.82) |
| **Awareness and threat perception about ticks and tick-borne diseases in humans** |  |
| **Awareness of TBDs in humans** |  |
| No/Do not know | 90 (31.58) |
| Yes | 195 (68.42) |
| **Worried about tick bite^†^** |  |
| Not worried | 71 (24.91) |
| Slightly - Somewhat worried | 117 (41.05) |
| Very worried | 93 (32.63) |
| **Prone to get a TBD^†§^** |  |
| No | 28 (14.36) |
| Yes | 164 (84.10) |
| **Probability to get a TBD^§^** |  |
| Not probable | 47 (24.10) |
| Slightly - Somewhat probable | 121 (62.05) |
| Highly probable | 27 (13.85) |
| **Worried about TBD^§^** |  |
| Not worried | 40 (20.51) |
| Slightly - Somewhat worried | 111 (56.92) |
| Very worried | 44 (22.56) |
| **Severity of TBD in humans^†§^** |  |
| Mild | 47 (24.10) |
| Moderate | 33 (16.92) |
| Severe - Very severe | 98 (50.26) |
| **Self-reported measures used to prevent/control ticks** |  |
| **Take dog(s) to the vet regularly^†^** |  |
| No dogs | 157 (55.09) |
| At least one dog | 125 (43.86) |
| **Spayed/neutered dog(s) older than 6 months^†ℨ^** |  |
| No dogs | 207 (78.41) |
| At least one dog | 55 (20.83) |
| **Home spraying with a "substance against ticks"** |  |
| No | 226 (79.30) |
| Yes | 59 (20.70) |
| **Application of acaricide sprays on dogs** |  |
| No dogs | 198 (69.47) |
| At least one dog | 87 (30.53) |
| **Administration or oral acaricides to dogs older than 2.5 months^†ℨ^** |  |
| No dogs | 248 (88.57) |
| At least one dog | 30 (10.71) |

**Table 1. Characteristics of dog owners living in Iquitos, Peru, awareness levels of tick-borne diseases and self-reported vector control methods**.

†Missing responses: Education level (1, 0.35%), Home income (28, 9.82), Worried about tick bite (4, 1.40%), prone to get a TBD (3, 1.54), severity of TBD (17, 8.27%), Take dogs to the vet regularly (3, 1.05%), spayed/neutered dog(s) (2, 0.76), administration of oral acaricides (2, 0.71%)

*Services and Customer Support (housewife, domestic worker, people working from home, cooker, food vendor, waiter/waitress, office worker, customer service); Primary Sector (agriculture, fishing, forestry); Secondary Sector (Construction); Tertiary Sector (transportation, merchant/trader); Other professionals (education and security).

§Question only asked to those who knew ticks can transmit diseases to humans (N = 195)

ℨPercentages may not sum up to 100% due to observations that do not apply do to age restrictions: Spayed/neutered dog(s) (21, 7.37%), administration of oral acaricides (5, 1.75%)
