## Supplementary material for "How threat perception of tick-borne diseases shapes preventive and control measures among dog owners in Iquitos, Peru": Table 2

**Table 2. Simple regressions between awareness of TBDs and threat perception for humans with self-reported measures to prevent/control ticks**

|  | **Self-reported measures to prevent/control ticks** | | | | | | | | | | | | | | |
| --- | --- | --- | --- | --- | --- | --- | --- | --- | --- | --- | --- | --- | --- | --- | --- |
|  | **Taken to veterinary**  **regularly**† | | | **Neuter/Spay dog(s)**  **(> 6 months)**† | | | **Home spray** | | | **Spray acaricides used**  **in dog(s)** | | | **Oral acaricides administered**  **to dog(s) (> 2.5 months)**† | | |
|  | **PR** | **95% CI** | **p** | **PR** | **95% CI** | **p** | **PR** | **95% CI** | **p** | **PR** | **95% CI** | **p** | **PR** | **95% CI** | **p** |
| **Awareness of TBDs in humans** |  |  |  |  |  |  |  |  |  |  |  |  |  |  |  |
| No/Do not know | Ref. | - | - | Ref. | - | - | Ref. | - | - | Ref. | - | - | Ref. | - | - |
| Yes | 1.40 | 1.02 - 1.92 | **0.040** | 1.85 | 1.01 - 3.41 | **0.047** | 2.01 | 1.10 - 3.69 | **0.024** | 1.21 | 0.81 - 1.81 | 0.346 | 1.50 | 0.67 - 3.36 | 0.328 |
| **Worried about tick bite†** |  |  |  |  |  |  |  |  |  |  |  |  |  |  |  |
| Not worried | Ref. | - | - | Ref. | - | - | Ref. | - | - | Ref. | - | - | Ref. | - | - |
| Slightly - Somewhat worried | 0.97 | 0.71 - 1.34 | 0.867 | 0.64 | 0.36 - 1.14 | 0.130 | 0.85 | 0.47 - 1.54 | 0.591 | 0.91 | 0.60 - 1.39 | 0.664 | 1.30 | 0.47 - 3.59 | 0.612 |
| Very worried | 0.89 | 0.63 - 1.26 | 0.508 | 0.82 | 0.46 - 1.45 | 0.491 | 1.12 | 0.63 - 2.00 | 0.702 | 0.76 | 0.47 - 1.23 | 0.265 | 1.94 | 0.73 - 5.20 | 0.186 |
| **Only participants who said yes to "ticks can transmit diseases to humans" were asked the following questions** | | | | | | | | | | | | | | | |
| **Threat perception**  **for humans** | **Taken to veterinary**  **frequently**† | | | **Neuter/Spay dog(s)**  **(> 6 months)**† | | | **Home spray** | | | **Spray acaricides used**  **in dog(s)** | | | **Oral acaricides administered**  **to dog(s) (> 2.5 months)**† | | |
|  | **PR** | **95% CI** | **p** | **PR** | **95% CI** | **p** | **PR** | **95% CI** | **p** | **PR** | **95% CI** | **p** | **PR** | **95% CI** | **p** |
| **Prone to get a TBD**†§ |  |  |  |  |  |  |  |  |  |  |  |  |  |  |  |
| No | Ref. | - | - | Ref. | - | - | Ref. | - | - | Ref. | - | - | Ref. | - | - |
| Yes | 1.08 | 0.70 - 1.65 | 0.734 | 1.11 | 0.52 - 2.38 | 0.788 | 2.50 | 0.83 - 7.53 | 0.102 | 1.59 | 0.76 - 3.35 | 0.219 | 0.57 | 0.29 - 1.42 | 0.228 |
| **Probability to get a TBD**§ |  |  |  |  |  |  |  |  |  |  |  |  |  |  |  |
| Not probable | Ref. | - | - | Ref. | - | - | Ref. | - | - | Ref. | - | - | Ref. | - | - |
| Slightly - Somewhat probable | 1.03 | 0.73 - 1.45 | 0.864 | 1.62 | 0.77 - 3.41 | 0.203 | 1.25 | 0.64 - 2.44 | 0.511 | 1.48 | 0.84 - 2.63 | 0.178 | 0.50 | 0.23 - 1.12 | 0.092 |
| Highly probable | 0.83 | 0.48 - 1.43 | 0.507 | 1.62 | 0.64 - 4.09 | 0.312 | 1.93 | 0.90 - 4.17 | 0.092 | 1.58 | 0.77 - 3.24 | 0.209 | 0.37 | 0.09 - 1.59 | 0.182 |
| **Worried about TBD**§ |  |  |  |  |  |  |  |  |  |  |  |  |  |  |  |
| Not worried | Ref. | - | - | Ref. | - | - | Ref. | - | - | Ref. | - | - | Ref. | - | - |
| Slightly - Somewhat worried | 1.22 | 0.81 - 1.83 | 0.336 | 1.09 | 0.54 - 2.21 | 0.815 | 1.26 | 0.63 - 2.54 | 0.515 | 1.59 | 0.88 - 2.85 | 0.123 | 0.56 | 0.23 - 1.35 | 0.198 |
| Very worried | 1.09 | 0.67 - 1.78 | 0.715 | 1.39 | 0.64 - 3.02 | 0.409 | 1.36 | 0.62 - 2.997 | 0.440 | 0.82 | 0.37 - 1.81 | 0.620 | 0.65 | 0.22 - 1.88 | 0.425 |
| **Severity of TBD in humans**†§ |  |  |  |  |  |  |  |  |  |  |  |  |  |  |  |
| Mild | Ref. | - | - | Ref. | - | - | Ref. | - | - | Ref. | - | - | Ref. | - | - |
| Moderate | 1.80 | 1.08 - 3.01 | **0.024** | 1.55 | 0.63 - 3.83 | 0.343 | 1.04 | 0.47 - 2.30 | 0.931 | 1.86 | 1.05 - 3.29 | **0.033** | 5.33 | 0.62 - 45.81 | 0.127 |
| Severe - Very severe | 1.62 | 1.02 - 2.56 | **0.042** | 1.73 | 0.82 - 3.68 | 0.152 | 1.09 | 0.59 - 2.03 | 0.785 | 1.03 | 0.59 - 1.81 | 0.910 | 6.80 | 0.92 - 50.20 | 0.060 |
| Prevalence ratios (PR), 95% Confidence intervals (95% CI) and p-values (p) calculated with generalized linear models with binomial family, link log, and robust variance | | | | | | | | | | | | | | | |
| †Percentages may not sum up to 100% due to missing responses | | | | | | | | | | | | | | | |
| **Those who knew ticks can transmit diseases to humans | | | | | | | | | | | | | | | |
| §Question only asked to those who knew ticks can transmit diseases to humans | | | | | | | | | | | | | | | |
