## Supplementary material for "How threat perception of tick-borne diseases shapes preventive and control measures among dog owners in Iquitos, Peru": Table 3

**Table 3. Association between awareness of TBDs or perceived severity for humans and self-reported measures to prevent/control ticks using a multiple regression analysis adjusted for socioeconomic factors.**

|  | **Self-reported measures to prevent/control ticks** | | | | | | | | | | | | | | |
| --- | --- | --- | --- | --- | --- | --- | --- | --- | --- | --- | --- | --- | --- | --- | --- |
|  | **Taken to the veterinary**  **regularly**** | | | **Neuter/Spay dog(s)**  **(>6 months)**** | | | **Home spray**** | | | **Spray acaricide used**  **in dog(s)**** | | | **Oral acaricides administered**  **to dogs(s) ( > 2.5 months)**** | | |
|  | **aPR** | **95% CI** | **p** | **aPR** | **95% CI** | **p** | **aPR** | **95% CI** | **p** | **aPR** | **95% CI** | **p** | **aPR** | **95% CI** | **p** |
| **Humans can get TBDs**† |  |  |  |  |  |  |  |  |  |  |  |  |  |  |  |
| No | Ref. | - | - | Ref. | - | - | Ref. | - | - | Ref. | - | - | Ref. | - | - |
| Yes | 1.10 | 0.80 - 1.50 | 0.555 | 1.55 | 0.85 - 2.83 | 0.153 | 1.87 | 1.01 - 3.47 | **0.048** | 1.07 | 0.70 - 1.62 | 0.764 | 1.23 | 0.49 - 3.08 | 0.658 |
| **Severity of TBD in humans**§† |  |  |  |  |  |  |  |  |  |  |  |  |  |  |  |
| Mild | Ref. | - | - | Ref. | - | - | Ref. | - | - | Ref. | - | - | Ref. | - | - |
| Moderate | 1.40 | 0.84 - 2.36 | 0.200 | 1.20 | 0.47 - 3.10 | 0.702 | 1.16 | 0.53 - 2.55 | 0.715 | 1.75 | 0.98 - 3.11 | **0.057** | 6.22 | 0.66 - 58.56 | 0.110 |
| Severe - Very severe | 1.36 | 0.86 - 2.14 | 0.188 | 1.33 | 0.61 - 2.90 | 0.467 | 0.98 | 0.52 - 1.85 | 0.955 | 0.92 | 0.52 - 1.62 | 0.761 | 7.31 | 1.08 - 49.36 | **0.041** |
| **Due to missing responses or age restrictions (neuter/spay and pills): N taken to veterinary regularly = 192, N neuter/spay dog = 178, N house spray = 194, N spray used on dogs = 194, N pills used on dogs = 190 | | | | | | | |  |  |  |  |  |  |  |  |
| *Prevalence ratios (PR), uPR = unadjusted, aPR = adjusted. 95% Confidence intervals (95% CI) and p-values (p) calculated with generalized linear models with Poisson family, link log, and robust variance | | | | | | | |  |  |  |  |  |  |  |  |
| §Question only asked to those who knew ticks can transmit diseases to humans | | | | | | | |  |  |  |  |  |  |  |  |
| †Adjusted for sex, age, education level, and home income | | | | | | | |  |  |  |  |  |  |  |  |
