## Supplementary material for "How threat perception of tick-borne diseases shapes preventive and control measures among dog owners in Iquitos, Peru": Table 4

**Table 4. Associations between sociodemographic factors and self-reported measures to prevent/control ticks using simple and multiple regression analyses.**

|  | **Self-reported measures to prevent/control ticks** | | | | | | | | | | | | | | | | | | | | | | | | |
| --- | --- | --- | --- | --- | --- | --- | --- | --- | --- | --- | --- | --- | --- | --- | --- | --- | --- | --- | --- | --- | --- | --- | --- | --- | --- |
|  | **Taken to the veterinary regularly**** | | | | | **Neuter/Spay dog (dogs >6 months)**** | | | | | **Home spray**** | | | | | **Acaricide spray used in dog(s)**** | | | | | **Oral acaricide administered to dog(s) (> 2.5 months)**** | | | | |
|  | **uPR** | **p** | **aPR** | **95% CI** | **p** | **uPR** | **p** | **aPR** | **95% CI** | **p** | **uPR** | **p** | **aPR** | **95% CI** | **p** | **uPR** | **p** | **aPR** | **95% CI** | **p** | **uPR** | **p** | **aPR** | **95% CI** | **p** |
| **Sex** |  |  |  |  |  |  |  |  |  |  |  |  |  |  |  |  |  |  |  |  |  |  |  |  |  |
| Female | Ref. | - | Ref. | - | - | Ref. | - | Ref. | - | - | Ref. | - | Ref. | - | - | Ref. | - | Ref. | - | - | Ref. | - | Ref. | - | - |
| Male | 0.93 | 0.64 | 0.73 | 0.54-0.99 | **0.040** | 1.25 | 0.37 | 1.17 | 0.70-1.94 | 0.56 | 0.69 | 0.19 | 0.63 | 0.36-1.10 | 0.10 | 0.90 | 0.59 | 0.82 | 0.54-1.24 | 0.34 | 1.24 | 0.55 | 1.09 | 0.50-2.37 | 0.82 |
| **Age (years)** |  |  |  |  |  |  |  |  |  |  |  |  |  |  |  |  |  |  |  |  |  |  |  |  |  |
| 18 - 43 | Ref. | - | Ref. | - | - | Ref. | - | Ref. | - | - | Ref. | - | Ref. | - | - | Ref. | - | Ref. | - | - | Ref. | - | Ref. | - | - |
| 44 - 59 | 0.68 | **0.032** | 0.88 | 0.62-1.22 | 0.45 | 1.06 | 0.83 | 1.23 | 0.74-2.05 | 0.42 | 1.19 | 0.50 | 1.41 | 0.84-2.37 | 0.19 | 1.01 | 0.97 | 1.13 | 0.76-1.68 | 0.56 | 0.64 | 0.32 | 0.67 | 0.28-1.62 | 0.37 |
| 60 - 80 | 1.06 | 0.71 | 1.73 | 1.21-2.49 | **0.003** | 0.68 | 0.31 | 0.79 | 0.35-1.77 | 0.57 | 0.92 | 0.80 | 1.20 | 0.56-2.55 | 0.64 | 0.70 | 0.21 | 0.82 | 0.43-1.54 | 0.53 | 1.02 | 0.9 | 1.00 | 0.38-2.62 | 0.996 |
| **Education level**† |  |  |  |  |  |  |  |  |  |  |  |  |  |  |  |  |  |  |  |  |  |  |  |  |  |
| No education - Less than high school | Ref. | - | Ref. | - | - | Ref. | - | Ref. | - | - | Ref. | - | Ref. | - | - | Ref. | - | Ref. | - | - | Ref. | - | Ref. | - | - |
| High school | 1.33 | 0.40 | 1.59 | 0.83-3.07 | 0.196 | 2.13 | 0.20 | 1.897 | 0.56-6.42 | 0.30 | 0.83 | 0.68 | 1.03 | 0.41-2.59 | 0.96 | 0.78 | 0.47 | 0.74 | 0.35-1.58 | 0.44 | 0.75 | 0.66 | 0.67 | 0.16-2.77 | 0.58 |
| Technical education | 2.18 | **0.018** | 2.83 | 1.43-5.60 | **0.003** | 1.70 | 0.39 | 1.37 | 0.38-4.91 | 0.63 | 1.18 | 0.70 | 1.60 | 0.58-4.36 | 0.36 | 1.47 | 0.23 | 1.40 | 0.65-2.98 | 0.39 | 1.48 | 0.53 | 1.08 | 0.26-4.46 | 0.92 |
| University - Grad school | 2.74 | **0.022** | 3.47 | 1.78-6.76 | **<0.001** | 3.50 | **0.030** | 2.87 | 0.85-9.70 | 0.09 | 1.71 | 0.19 | 2.41 | 0.89-6.51 | 0.08 | 1.44 | 0.26 | 1.39 | 0.64-3.02 | 0.40 | 1.46 | 0.54 | 0.96 | 0.25-3.66 | 0.96 |
| **House income** |  |  |  |  |  |  |  |  |  |  |  |  |  |  |  |  |  |  |  |  |  |  |  |  |  |
| ≤ 1,000 | Ref. | - | Ref. | - | - | Ref. | - | Ref. | - | - | Ref. | - | Ref. | - | - | Ref. | - | Ref. | - | - | Ref. | - | Ref. | - | - |
| 1,001 - 2,500 | 1.47 | 0.11 | 1.2 | 0.76-1.89 | 0.44 | 1.30 | 0.51 | 1.05 | 0.50-2.25 | 0.897 | 0.84 | 0.57 | 0.73 | 0.39-1.38 | 0.33 | 1.06 | 0.83 | 0.96 | 0.55-1.69 | 0.898 | 1.11 | 0.86 | 1.03 | 0.34-3.10 | 0.96 |
| > 2,500 | 1.86 | **0.010** | 1.26 | 0.78-2.04 | 0.34 | 1.78 | 0.14 | 1.32 | 0.60-2.94 | 0.49 | 1.02 | 0.95 | 0.81 | 0.39-1.66 | 0.56 | 1.31 | 0.32 | 1.09 | 0.60-1.97 | 0.78 | 2.30 | 0.12 | 1.94 | 0.58-6.53 | 0.28 |
| **Due to missing responses or age restrictions (neuter/spay and pills): N taken to veterinary regularly = 281, N neuter/spay dog = 261, N house spray = 284, N spray used on dogs = 284, N pills used on dogs = 277.  * Prevalence ratios (PR), uPR = unadjusted, aPR = adjusted. 95% Confidence intervals (95% CI) and p-values (p) calculated with generalized linear models with Poisson family, link log, and robust variance. †Observations excluded due to missing responses (<2% of observations) | | | | | | | | | | | | | | | | | | | | | | | | | |
