## Supplementary material for "How threat perception of tick-borne diseases shapes preventive and control measures among dog owners in Iquitos, Peru": Table 5

**Table 5. Associations between sociodemographic factors and awareness of TBDs or threat perception using simple regression analyses.**

|  | **Threat perception for humanas** | | | | | | | | | | | | | | | | | | | | | | | | | | | | | | |
| --- | --- | --- | --- | --- | --- | --- | --- | --- | --- | --- | --- | --- | --- | --- | --- | --- | --- | --- | --- | --- | --- | --- | --- | --- | --- | --- | --- | --- | --- | --- | --- |
| **Socioeconomic factors** | **People can get TBD** | | | **Worried about tick bite** | | | | | | |  |  | |  | |  |  | |  | |  |  | |  | |  |  | | |  | |
|  | **OR*** | **95% CI** | **p** | **Slightly - Somewhat**  **worried** | | | **Very worried** | | | |  |  | |  | |  |  | |  | |  |  | |  | |  |  | | |  | |
|  |  |  |  | **OR** | **95% CI** | **p** | **OR** | **95% CI** | **p** | |  |  | |  | |  |  | |  | |  |  | |  | |  |  | | |  | |
| **Sex** |  |  |  |  |  |  |  |  |  | |  |  | |  | |  |  | |  | |  |  | |  | |  |  | | |  | |
| Female | Ref. | - | - | Ref. | - | - | Ref. | - | - | |  |  | |  | |  |  | |  | |  |  | |  | |  |  | | |  | |
| Male | 1.17 | 0.68-2.03 | 0.563 | 0.31 | 0.17-0.58 | **<0.001** | 0.19 | 0.10-0.39 | **<0.001** | |  |  | |  | |  |  | |  | |  |  | |  | |  |  | | |  | |
| **Age (years)** |  |  |  |  |  |  |  |  |  | |  |  | |  | |  |  | |  | |  |  | |  | |  |  | | |  | |
| 18 - 43 | Ref. | - | - | Ref. | - | - | Ref. | - | - | |  |  | |  | |  |  | |  | |  |  | |  | |  |  | | |  | |
| 44 - 59 | 0.42 | 0.23-0.76 | **0.004** | 0.34 | 0.17-0.70 | **0.003** | 0.58 | 0.29-1.17 | 0.129 | |  |  | |  | |  |  | |  | |  |  | |  | |  |  | | |  | |
| 60 - 80 | 0.30 | 0.16-0.60 | **0.001** | 0.40 | 0.18-0.87 | **0.022** | 0.41 | 0.17-0.97 | **0.042** | |  |  | |  | |  |  | |  | |  |  | |  | |  |  | | |  | |
| **Education level**† |  |  |  |  |  |  |  |  |  | |  |  | |  | |  |  | |  | |  |  | |  | |  |  | | |  | |
| No education - Elementary school | Ref. | - | - | Ref. | - | - | Ref. | - | - | |  |  | |  | |  |  | |  | |  |  | |  | |  |  | | |  | |
| High school | 4.04 | 1.74-9.36 | **0.001** | 1.09 | 0.42-2.79 | 0.861 | 1.58 | 0.57-4.31 | 0.380 | |  |  | |  | |  |  | |  | |  |  | |  | |  |  | | |  | |
| Technical education | 7.76 | 3.11-19.42 | **<0.001** | 1.43 | 0.52-3.93 | 0.491 | 2.14 | 0.73-6.26 | 0.165 | |  |  | |  | |  |  | |  | |  |  | |  | |  |  | | |  | |
| University - Grad school | 14.18 | 5.34-37.64 | **<0.001** | 2.31 | 0.85-6.27 | 0.099 | 1.63 | 0.54-4.93 | 0.387 | |  |  | |  | |  |  | |  | |  |  | |  | |  |  | | |  | |
| **House income** |  |  |  |  |  |  |  |  |  | |  |  | |  | |  |  | |  | |  |  | |  | |  |  | | |  | |
| ≤ 1,000 | Ref. | - | - | Ref. | - | - | Ref. | - | - | |  |  | |  | |  |  | |  | |  |  | |  | |  |  | | |  | |
| 1,001 - 2,500 | 1.16 | 0.59-2.27 | 0.664 | 1.87 | 0.84-4.17 | 0.126 | 2.04 | 0.85- 4.92 | 0.112 | |  |  | |  | |  |  | |  | |  |  | |  | |  |  | | |  | |
| > 2,500 | 2.50 | 1.16-5.41 | **0.020** | 1.49 | 0.64-3.47 | 0.359 | 1.896 | 0.76-4.76 | 0.173 | |  |  | |  | |  |  | |  | |  |  | |  | |  |  | | |  | |
| **Only participants who said yes to "ticks can transmit diseases to humans" were asked the following questions** | | | | | | | | | | | | | | | | | | | | | | | | | | | | | | | |
|  | **Prone to get TBD**§ | | | **Probability to get a TBD**§ | | | | | | | **Worried about TBDs**§ | | | | | | | | | | **Severity of TBD among humans**§ | | | | | | | | | | |
|  | **OR*** | **95% CI** | **p** | **Slightly - Somewhat**  **probable** | | | **Very probable** | | | | **Slightly - Somewhat**  **worried** | | | | | **Very worried** | | | | | **Moderate** | | | | | **Severe - Very severe** | | | | | |
|  |  |  |  | **OR** | **95% CI** | **p** | **OR** | **95% CI** | | **p** | **OR** | | **95% CI** | | **p** | **OR** | | **95% CI** | | **p** | **OR** | | **95% CI** | | **p** | **OR** | | **95% CI** | | | **p** |
| **Sex** |  |  |  |  |  |  |  |  | |  |  | |  | |  |  | |  | |  |  | |  | |  |  | |  | | |  |
| Female | Ref. | - | - | Ref. | - | - | Ref. | - | | - | Ref. | | - | | - | Ref. | | - | | - | Ref. | | - | | - | Ref. | | - | | | - |
| Male | 0.51 | 0.22-1.14 | 0.101 | 0.57 | 0.28-1.15 | 0.116 | 0.47 | 0.17-1.33 | | 0.156 | 0.63 | | 0.30-1.33 | | 0.229 | 0.23 | | 0.08-0.64 | | **0.005** | 1.47 | | 0.59 -3.64 | | 0.405 | 0.57 | | 0.27-1.21 | | | 0.146 |
| **Age (years)** |  |  |  |  |  |  |  |  | |  |  | |  | |  |  | |  | |  |  | |  | |  |  | |  | | |  |
| 18 - 43 | Ref. | - | - | Ref. | - | - | Ref. | - | | - | Ref. | | - | | - | Ref. | | - | | - | Ref. | | - | | - | Ref. | | - | | | - |
| 44 - 59 | 1.26 | 0.43-3.67 | 0.670 | 1.32 | 0.58-3.01 | 0.515 | 1.28 | 0.41-3.96 | | 0.673 | 0.84 | | 0.36-2.00 | | 0.700 | 0.85 | | 0.30-2.40 | | 0.754 | 0.43 | | 0.14 -1.37 | | 0.155 | 0.64 | | 0.29-1.40 | | | 0.263 |
| 60 - 80 | 0.42 | 0.15-1.15 | 0.091 | 1.20 | 0.43-3.34 | 0.723 | 0.91 | 0.20-4.11 | | 0.904 | 0.38 | | 0.13-1.07 | | 0.067 | 0.85 | | 0.27-2.63 | | 0.773 | 3.26 | | 0.58-18.34 | | 0.180 | 2.50 | | 0.52-11.98 | | | 0.252 |
| **Education level**† |  |  |  |  |  |  |  |  | |  |  | |  | |  |  | |  | |  |  | |  | |  |  | |  | | |  |
| No education - Elementary school | Ref. | - | - | Ref. | - | - | Ref. | - | | - | Ref. | | - | | - | Ref. | | - | | - | Ref. | | - | | - | Ref. | | - | | | - |
| High school | 4.75 | 0.94-24.03 | 0.060 | 7.19 | 1.58-32.67 | **0.011** | 6.25 | 0.60-64.86 | | 0.125 | 5.63 | | 1.07-29.73 | | **0.042** | 2.52 | | 0.46-13.80 | | 0.287 | ** | | ** | | ** | 2.00 | | 0.40-10.03 | | | 0.399 |
| Technical education | 2.94 | 0.61-14.20 | 0.180 | 2.13 | 0.50-0.04 | 0.304 | 2.65 | 0.27-26.24 | | 0.406 | 5.04 | | 0.95-26.69 | | 0.057 | 1.78 | | 0.32-10.01 | | 0.514 | ** | | ** | | ** | 2.20 | | 0.44-11.03 | | | 0.339 |
| University - Grad school | 2.7 | 0.58-12.61 | 0.207 | 3.28 | 0.78-13.78 | 0.105 | 2.19 | 0.21-22.34 | | 0.509 | 2.82 | | 0.57-14.04 | | 0.205 | 0.94 | | 0.18-4.997 | | 0.943 | ** | | ** | | ** | 6.67 | | 1.25-35.74 | | | **0.027** |
| **House income** |  |  |  |  |  |  |  |  | |  |  | |  | |  |  | |  | |  |  | |  | |  |  | |  | | |  |
| No reported | 0.75 | 0.15-3.82 | 0.726 | 1.74 | 0.39-7.81 | 0.472 | 2.25 | 0.31-16.41 | | 0.424 | 1.46 | | 0.36-5.91 | | 0.599 | 0.75 | | 0.10-5.47 | | 0.777 | 3.06 | | 0.53-17.46 | | 0.209 | 0.65 | | 0.16-2.72 | | | 0.560 |
| ≤ 1,000 | Ref. | - | - | Ref. | - | - | Ref. | - | | - | Ref. | | - | | - | Ref. | | - | | - | Ref. | | - | | - | Ref. | | - | | | - |
| 1,001 - 2,500 | 1.62 | 0.44-6.02 | 0.469 | 1.61 | 0.60-4.29 | 0.341 | 1.80 | 0.44-7.31 | | 0.411 | **3.38** | | 1.12-10.13 | | **0.030** | 3.56 | | 0.95-13.37 | | 0.060 | 2.65 | | 0.61-11.43 | | 0.192 | 1.75 | | 0.66-4.59 | | | 0.258 |
| > 2,500 | 0.62 | 0.18-2.07 | 0.434 | 0.95 | 0.36-2.47 | 0.912 | 0.90 | 0.21-3.78 | | 0.886 | 0.89 | | 0.33-2.39 | | 0.820 | 1.34 | | 0.40-4.56 | | 0.637 | 3.67 | | 0.81-16.54 | | 0.091 | 2.55 | | 0.92-7.09 | | | 0.072 |
| Odds ratios (OR), 95% Confidence intervals (95% CI) and p-values (p) calculated with multinomial logistic regression. Reference categories for outcomes are not worried (for worried about tick bites and TBs), not prone (for prone to get TBDs), not probable (for probability to get TBDs), and mild (for severity of TBDs). | | | | | | | | | | | | | | | | | | | | | | | | | | | | | | | |
| OR* calculated with logistic regression due to the nature of the outcome variable | | | | | | | | | | | | | | | | | | | | | | | | | | | | |  | | |
| †Percentages may not sum up to 100% due to missing values | | |  |  |  |  |  |  |  | |  |  | |  | |  |  | |  | |  |  | |  | |  |  | | |  | |
| **Those who knew ticks can transmit diseases to humans | | |  |  |  |  |  |  |  | |  |  | |  | |  |  | |  | |  |  | |  | |  |  | | |  | |
| §Question only asked to those who knew ticks can transmit diseases to humans | | | | | | | | | | | | | | | | | | | | | | | | | | | | | | | |
