## Supplementary Table 1 for "How threat perception of tick-borne diseases shapes preventive and control measures among dog owners in Iquitos, Peru"

**Sup. Table 1. Type of products used by the participants to treat their house against arthropods.**

|  | **N (%)** |
| --- | --- |
| **Type** |  |
| Commercially available insecticides^1^ | 25 (42.37%) |
| Phenol-based products^2^ | 11 (18.64%) |
| Endectocides and ectoparasiticides^3^ | 5 (8.47%) |
| Disinfectants^4^ | 5 (8.47%) |
| Did not recall name | 13 (22.03%) |

^1^Include pyrethroids and phenylpyrazoles

^2^Include Creoline^®^, and hydrocarbons as the petroleum, sometimes mixed with bleach

^3^Include ivermectin and amitraz, and others

^4^Mainly bleach
