## Supplementary Table 2 for "How threat perception of tick-borne diseases shapes preventive and control measures among dog owners in Iquitos, Peru"

**Sup. Table 2. Associations between awareness of TBDs or threat perception for humans and self-reported measures used to prevent/control ticks using a bivariate analysis.**

|  | **Self-reported practices to prevent/control ticks** | | | | | | | | | |
| --- | --- | --- | --- | --- | --- | --- | --- | --- | --- | --- |
| **Threat perception**  **for humans** | **Taken to veterinary**  **regularly**† | | **Neuter/Spay dog(s)**  **(> 6 months)**† | | **Home spray** | | **Spray acaricide used in dog(s)** | | **Oral acaricides administered**  **to dog(s) (> 2.5 months)**† | |
|  | **At least one dog**  125 (43.86) | **p** | **At least one dog**  55 (20.83) | **p** | **Yes**  59 (20.70) | **p** | **Used in one/all**  **inc. or corr. frequency**  87 (30.53) | **p** | **Used in one/all**  **inc. or corr. frequency**  30 (10.71) | **p** |
| **Awareness of TBDs in humans** |  | **0.029** |  | **0.036** |  | **0.016** |  | 0.336 |  | 0.319 |
| No/Do not know | 31 (34.83) |  | 11 (13.25) |  | 11 (12.22) |  | 24 (26.67) |  | 8 (8.05) |  |
| Yes | 94 (48.70) |  | 44 (24.58) |  | 48 (24.62) |  | 63 (32.31) |  | 23 (12.04) |  |
| **Worried about tick bite**† |  | 0.775 |  | 0.314 |  | 0.593 |  | 0.522 |  | 0.376¶ |
| Not worried | 33 (46.48) |  | 17 (26.98) |  | 15 (21.13) |  | 24 (33.80) |  | 5 (7.35) |  |
| Slightly - Somewhat worried | 52 (45.22) |  | 19 (17.27) |  | 21 (17.95) |  | 36 (30.77) |  | 11 (9.57) |  |
| Very worried | 38 (41.30) |  | 19 (22.09) |  | 22 (23.66) |  | 24 (25.81) |  | 13 (14.29) |  |
| **Only participants who said yes to "ticks can transmit diseases to humans" were asked the following questions** | | | | | | | | | | |
| **Threat perception**  **for humans** | **Taken to veterinary**  **frequently**† | | **Neuter/Spay dog(s)**  **(> 6 months)**† | | **Home spray** | | **Spray acaricide used in dog(s)** | | **Oral acaricides administered**  **to dog(s) (> 2.5 months)**† | |
|  | **At least one dog**  94 (48.21) | **p** | **At least one dog**  44 (24.44) | **p** | **Yes**  48 (24.62) | **p** | **Used in one/all**  **inc. or corr. frequency**  63 (32.31) | **p** | **Used in one/all**  **inc. or corr. frequency**  23 (11.98) | **p** |
| **Prone to get a TBD**†§ |  | 0.727 |  | 0.785 |  | 0.094¶ |  | 0.183 |  | 0.327¶ |
| No | 13 (46.43) |  | 6 (22.22) |  | 3 (10.71) |  | 6 (21.43) |  | 5 (18.52) |  |
| Yes | 81 (50.00) |  | 37 (24.67) |  | 44 (26.83) |  | 56 (34.15) |  | 17 (10.56) |  |
| **Probability to get a TBD**§ |  | 0.661 |  | 0.396 |  | 0.220 |  | 0.317 |  | 0.193¶ |
| Not probable | 23 (48.94) |  | 7 (16.67) |  | 9 (19.15) |  | 11 (23.40) |  | 9 (20.00) |  |
| Slightly - Somewhat probable | 60 (50.42) |  | 30 (27.03) |  | 29 (23.97) |  | 42 (34.71) |  | 12 (10.08) |  |
| Highly probable | 11 (40.74) |  | 7 (26.92) |  | 10 (37.04) |  | 10 (37.04) |  | 2 (7.41) |  |
| **Worried about TBD**§ |  | 0.570 |  | 0.648 |  | 0.723 |  | **0.038** |  | 0.401¶ |
| Not worried | 17 (42.50) |  | 8 (21.62) |  | 8 (20.00) |  | 10 (25.00) |  | 7 (17.95) |  |
| Slightly - Somewhat worried | 57 (51.82) |  | 24 (23.53) |  | 28 (25.23) |  | 44 (39.64) |  | 11 (10.09) |  |
| Very worried | 20 (46.51) |  | 12 (30.00) |  | 12 (27.27) |  | 9 (20.45) |  | 5 (11.63) |  |
| **Severity of TBD in humans**†§ |  | **0.038** |  | 0.316 |  | 0.960 |  | **0.036** |  | **0.060**¶ |
| Mild | 15 (31.91) |  | 7 (16.67) |  | 11 (23.40) |  | 13 (27.66) |  | 1 (2.27) |  |
| Moderate | 19 (57.58) |  | 8 (25.81) |  | 8 (24.24) |  | 17 (51.52) |  | 4 (12.12) |  |
| Severe - Very severe | 50 (51.55) |  | 26 (28.89) |  | 25 (25.51) |  | 28 (28.57) |  | 15 (15.46) |  |
| †Percentages may not sum up to 100% due to missing responses | | | | | | | | | | |
| **Those who knew ticks can transmit diseases to humans | | | | | | | | | | |
| §Question only asked to those who knew ticks can transmit diseases to humans | | | | | | | | | | |
| ¶p-values calculated using Fisher's exact test | | | | | | | | | | |
