## Supplementary Table 3 for "How threat perception of tick-borne diseases shapes preventive and control measures among dog owners in Iquitos, Peru"

| **Sup. Table 3. Associations between sociodemographic factors and self-reported measures to prevent/control ticks using bivariate analyses.** | | | | | | | | | | |
| --- | --- | --- | --- | --- | --- | --- | --- | --- | --- | --- |
|  | **Self-reported measures to prevent/control ticks** | | | | | | | | | |
| **Sociodemographic factors** | **Taken to veterinarian**  **regularly**† | | **Neuter/Spay dog(s)**  **(> 6 months)**† | | **Home spray** | | **Spray acaricide used**  **in dog(s)** | | **Oral acaricide administered**  **to dog(s) (> 2.5 months)**† | |
|  | **At least one dog**  125 (43.86) | **p** | **At least one dog**  55 (20.83) | **p** | **Yes**  59 (20.70) | **p** | **Used in one/all**  **inc. or corr. frequency**  87 (30.53) | **p** | **Used in one/all**  **inc. or corr. frequency**  30 (10.71) | **p** |
| **Sex** |  | 0.708 |  | 0.401 |  | 0.163 |  | 0.547 |  | 0.563 |
| Female | 87 (45.08) |  | 35 (19.55) |  | 45 (22.96) |  | 62 (31.63) |  | 19 (10.05) |  |
| Male | 38 (42.70) |  | 20 (24.10) |  | 14 (15.73) |  | 25 (28.09) |  | 11 (12.36) |  |
| **Age (years)** |  | **0.037** |  | 0.487 |  | 0.697 |  | 0.353 |  | 0.588 |
| 18 - 43 | 74 (48.05) |  | 31 (21.99) |  | 31 (20.00) |  | 50 (32.26) |  | 18 (12.00) |  |
| 44 - 59 | 26 (32.05) |  | 17 (23.29) |  | 19 (23.75) |  | 26 (32.50) |  | 6 (7.69) |  |
| 60 - 80 | 25 (25.08) |  | 7 (14.58) |  | 9 (18.00) |  | 11 (22.00) |  | 6 (12.00) |  |
| **Education level**† |  | **<0.001** |  | **0.027¶** |  | 0.084 |  | **0.024** |  | 0.387¶ |
| No education - Less than high school | 8 (23.53) |  | 3 (9.38) |  | 6 (17.65) |  | 9 (26.47) |  | 3 (9.38) |  |
| High school | 32 (31.37) |  | 18 (20.00) |  | 15 (14.71) |  | 21 (20.59) |  | 7 (7.00) |  |
| Technical education | 37 (51.39) |  | 11 (15.94) |  | 15 (20.83) |  | 28 (38.89) |  | 10 (13.89) |  |
| University - Grad school | 47 (64.38) |  | 23 (32.86) |  | 23 (30.26) |  | 29 (38.16) |  | 10 (13.70) |  |
| **House income**† |  | **0.018** |  | 0.241 |  | 0.715 |  | 0.472 |  | 0.095¶ |
| ≤ 1,000 | 15 (28.85) |  | 7 (15.22) |  | 12 (23.08) |  | 14 (26.92) |  | 4 (7.84) |  |
| 1,001 - 2,500 | 51 (42.86) |  | 21 (19.63) |  | 23 (19.17) |  | 34 (28.33) |  | 10 (8.62) |  |
| > 2,500 | 45 (53.57) |  | 22 (27.16) |  | 20 (23.53) |  | 30 (35.29) |  | 15 (18.07) |  |
| †Percentages may not sum up to 100% due to missing values | | | | | | | | | | |
| **Those who knew ticks can transmit diseases to humans | | | | | | | | | | |
| §Question only asked to those who knew ticks can transmit diseases to humans | | | | | | | | | | |
| ¶p-values calculated using Fisher's exact test | | | | | | | | | | |
