## Supplementary Table 4 for "How threat perception of tick-borne diseases shapes preventive and control measures among dog owners in Iquitos, Peru"

**Sup. Table 4a. Associations between sociodemographic factors and threat perception using bivariate analyses.**

| **Sociodemographic factors** | **Awareness of TBDs in**  **humans** | | **Worried about tick bite**† | | |
| --- | --- | --- | --- | --- | --- |
|  | **Yes**  **195 (68.42)** | **p** | **Slightly-Somewhat worried**  **117 (41.05)** | **Very worried**  93 (32.63) | **p** |
| **Sex** |  | 0.563 |  |  | **<0.001** |
| Female | 132 (67.35) |  | 86 (44.10) | 76 (38.97) |  |
| Male | 63 (70.79) |  | 31 (36.05) | 17 (19.77) |  |
| **Age (years)** |  | **<0.001** |  |  | **0.018** |
| 18 - 43 | 121 (78.06) |  | 75 (48.39) | 52 (33.55) |  |
| 44 - 59 | 48 (60.00) |  | 24 (30.77) | 28 (35.90) |  |
| 60 - 80 | 26 (52.00) |  | 18 (37.50) | 13 (27.08) |  |
| **Education level**† |  | **<0.001** |  |  | 0.199 |
| No education - Less than high school | 10 (29.41) |  | 13 (39.39) | 9 (27.27) |  |
| High school | 64 (62.75) |  | 36 (36.00) | 36 (36.00) |  |
| Technical education | 55 (76.39) |  | 27 (38.03) | 28 (39.44) |  |
| University - Grad school | 65 (85.53) |  | 41 (53.95) | 20 (26.32) |  |
| **House income**† |  | **0.029** |  |  | 0.461 |
| ≤ 1,000 | 32 (61.54) |  | 20 (40.00) | 13 (26.00) |  |
| 1,001 - 2,500 | 78 (65.00) |  | 55 (46.22) | 39 (32.77) |  |
| > 2,500 | 68 (80.00) |  | 35 (41.67) | 29 (34.52) |  |
