## Supplementary Table 4b for "How threat perception of tick-borne diseases shapes preventive and control measures among dog owners in Iquitos, Peru"

| **Sup. Table 4b. Associations between sociodemographic factors and threat perception using bivariate analyses in participants who said yes to "ticks can transmit diseases to humans".** | | | | | | | | | | | |
| --- | --- | --- | --- | --- | --- | --- | --- | --- | --- | --- | --- |
| **Sociodemographic factors** | **Prone to get a TBD**§ | | **Probability to get a TBD**§ | | | **Worried about TBD**§ | | | **Severity of TBD among humans**†§ | | |
|  | **Yes**  164 (84.10) | **p** | **Slightly-Somewhat probable**  121 (62.05) | **Highly probable**  27 (13.85) | **p** | **Slightly-Somewhat worried**  111 (56.92) | **Very worried**  55 (22.56) | **p** | **Moderate**  33 (16.92) | **Severe-Very severe**  98 (50.26) | **p** |
| **Sex** |  | 0.097 |  |  | 0.210 |  |  | **0.014** |  |  | **0.058** |
| Female | 114 (88.37) |  | 85 (64.39) | 20 (15.15) |  | 73 (55.30) | 37 (28.03) |  | 18 (14.75) | 74 (60.66) |  |
| Male | 50 (79.37) |  | 36 (57.14) | 7 (11.11) |  | 38 (60.32) | 7 (11.11) |  | 15 (26.79) | 24 (42.86) |  |
| **Age (years)** |  | 0.162 |  |  | 0.972 |  |  | 0.331 |  |  | 0.261 |
| 18 - 43 | 104 (86.67) |  | 73 (60.33) | 17 (14.05) |  | 73 (60.33) | 26 (21.49) |  | 23 (19.33) | 66 (55.46) |  |
| 44 - 59 | 41 (89.13) |  | 31 (64.58) | 7 (14.58) |  | 28 (58.33) | 10 (20.83) |  | 5 (12.20) | 21 (51.22) |  |
| 60 - 80 | 19 (73.08) |  | 17 (65.38) | 3 (11.54) |  | 10 (38.46) | 8 (30.77) |  | 5 (27.78) | 11 (61.11) |  |
| **Education level**† |  | 0.238¶ |  |  | 0.071¶ |  |  | 0.256¶ |  |  | **0.043¶** |
| No education - Less than high school | 6 (66.67) |  | 4 (40.00) | 1 (10.00) |  | 3 (30.00) | 3 (30.00) |  | 0 (0.00) | 3 (42.86) |  |
| High school | 57 (90.48) |  | 46 (71.88) | 10 (15.62) |  | 38 (59.38) | 3 (30.00) |  | 10 (18.18) | 27 (49.09) |  |
| Technical education | 47 (85.45) |  | 29 (52.73) | 9 (16.36) |  | 34 (61.82) | 12 (21.82) |  | 8 (15.09) | 28 (52.83) |  |
| University - Grad school | 54 (84.38) |  | 42 (64.62) | 7 (10.77) |  | 36 (55.38) | 12 (18.46) |  | 15 (23.81) | 40 (63.49) |  |
| **House income**† |  | 0.139¶ |  |  | 0.660¶ |  |  | 0.051 |  |  | 0.370¶ |
| ≤ 1,000 | 25 (86.21) |  | 19 (59.38) | 4 (12.50) |  | 17 (53.12) | 6 (18.75) |  | 3 (10.71) | 14 (50.00) |  |
| 1,001 - 2,500 | 71 (91.03) |  | 51 (65.38) | 12 (15.38) |  | 51 (65.38) | 19 (24.36) |  | 13 (18.31) | 40 (56.34) |  |
| > 2,500 | 54 (79.41) |  | 40 (58.82) | 8 (11.76) |  | 32 (47.06) | 17 (25.00) |  | 12 (19.05) | 39 (61.90) |  |
| †Percentages may not sum up to 100% due to missing values | | | | | | | | | | | |
| **Those who knew ticks can transmit diseases to humans | | | | | | | | | | | |
| §Question only asked to those who knew ticks can transmit diseases to humans | | | | | | | | | | | |
| ¶p-values calculated using Fisher's exact test | | | | | | | | | | | |
