## Supplementary Table 5 for "How threat perception of tick-borne diseases shapes preventive and control measures among dog owners in Iquitos, Peru"

**Sup. Table 5. Characteristics of individuals excluded from final analyses due to missing values for home income and/or perceived TBD severity.**

| **Characteristics** | **Home income** | | **p** | **TBD severity** | | **p** |
| --- | --- | --- | --- | --- | --- | --- |
|  | **No missing** | **Missing** |  | **No missing** | **Missing** |  |
|  | **(n= 257)** | **(n=28)** |  | **(n= 178)** | **(n=17)** |  |
| **Owner sex** |  |  | 0.333 |  |  | 0.413 |
| Female | 179 (91.3) | 17 (8.7) |  | 122 (92.4) | 10 (7.6) |  |
| Male | 78 (87.6) | 11 (12.4) |  | 56 (88.9) | 7 (11.1) |  |
| **Owner age** |  |  | 0.475 |  |  | **<0.001** |
| 18-43 | 137 (88.4) | 18 (11.6) |  | 119 (98.3) | 2 (1.7) |  |
| 44-59 | 73 (91.2) | 7(8.8) |  | 41 (85.4) | 7 (14.6) |  |
| 60-80 | 47 (94.0) | 3 (6.0) |  | 18 (69.2) | 8 (30.7) |  |
| **Owner education** |  |  | 0.589 |  |  | **<0.001** |
| No studies - elementary | 31 (91.2) | 3 (8.8) |  | 7 (70.0) | 3 (30.0) |  |
| High school | 89 (87.2) | 13 (12.8) |  | 55 (85.9) | 9 (14.1) |  |
| Institute | 65 (90.3) | 7 (9.7) |  | 53 (96.4) | 2 (3.6) |  |
| University - Graduate | 71 (93.4) | 5 (6.6) |  | 63 (96.9) | 2 (3.1) |  |
| **Dog frequently taken to vet** |  |  | 0.408 |  |  | 0.249 |
| No | 144 (91.7) | 13 (8.3) |  | 93 (93.9) | 6 (6.1) |  |
| At least one dog | 111 (88.8) | 14 (11.2) |  | 84 (89.4) | 10 (10.6) |  |
| **Dog reporductive status** |  |  | 0.261 |  |  | 0.812 |
| No dog older than 6 months | 134 (83.8) | 26 (16.3) |  | 122 (90.4) | 13 (9.6) |  |
| At least one dog older than 6 months | 50 (90.9) | 5 (9.1) |  | 41 (93.2) | 3 (6.8) |  |
| Does not apply | 21 (100.0) | 0 (0) |  | 14 (93.3) | 1 (6.7) |  |
| **Home spray** |  |  | 0.378 |  |  | 0.931 |
| No | 202 (89.4) | 24 (10.6) |  | 134 (91.2) | 13 (8.8) |  |
| Yes | 55 (93.2) | 4 (6.8) |  | 44 (91.7) | 15 (8.3) |  |
| **Spray acaricide used in dog(s)** |  |  | 0.845 |  |  | 0.789 |
| No spray used | 179 (90.4) | 19 (9.6) |  | 120 (90.9) | 12 (9.1) |  |
| Spray used in at least | 78 (89.7) | 9 (10.3) |  | 58 (92.1) | 5 (7.9) |  |
| **Oral acaricide administered to dog(s)** |  |  | 0.321 |  |  | 0.573^¶^ |
| No pill used in dog | 221 (89.1) | 27 (10.9) |  | 154 (91.7) | 14 (8.3) |  |
| Pill used in at least dog | 29 (96.7) | 1 (3.3) |  | 20 (86.9) | 3 (13.1) |  |
| Does not apply | 255 (100.0) | 0 (0) |  | 3 (100) | 0 (0) |  |

^¶^p-values calculated using Fisher's exact test
